# Multicentric validation of a Multimorbidity Adjusted Disability Score to stratify depression-related risks using temporal disease maps

**DOI:** 10.1101/2023.09.04.23295005

**Authors:** Rubèn González-Colom, Kangkana Mitra, Emili Vela, Andras Gezsi, Teemu Paajanen, Zsofia Gal, Gabor Hullam, Hannu Mäkinen, Tamas Nagy, Mikko Kuokkanen, Jordi Piera-Jiménez, Josep Roca, Peter Antal, Gabriella Juhasz, Isaac Cano

## Abstract

In the EU project TRAJECTOME, we used a novel methodology to identify temporal disease maps of depression and highly prevalent co-occurring disease conditions. This information was combined with disability weights established by the Global Burden of Disease Study 2019 to create a depression-related health risk assessment tool, the Multimorbidity Adjusted Disability Score (MADS). MADS was used to stratify over one million cases from three different cohorts and evaluate the impact on utilisation of healthcare resources, mortality, pharmacological burden, healthcare expenditure and multimorbidity progression. Results indicate statistically significant associations between MADS and increased mortality rate (P <.001), heightened healthcare utilization (i.e. emergency room visits P <.001; hospitalizations P <.001; pharmaceutical prescriptions P <.001; total healthcare expenditure P <.001), and a higher risk of disease progression and incidence of new depression-related comorbidities. MADS seems to be a promising approach to predict depression-related health risk and depression’s impact on individuals and healthcare systems, which can be tested in other diseases; nevertheless, clinical validation is still necessary.

## INTRODUCTION

The continuously increasing prevalence of multimorbidity is a pressing concern tied to complex clinical situations that can markedly impair patients’ quality of life and result in escalated healthcare costs^1,2^. It is widely accepted that diseases can frequently co-occur in specific patterns forming clusters. Particularly noteworthy is the cluster of Major Depressive Disorder (MDD) [F32 and F33 ICD-10-CM^3^] and other mental and somatic illnesses. Furthermore, it is recognised that individuals afflicted with MDD may encounter additional obstacles in effectively managing their overall health^4,5^, a feature that heightens the possibility that a disease-centred approach might lead to suboptimal management of patients with multiple, related and disabling chronic conditions^6^.

To evaluate patients’ clinical complexity and managing the impact of multimorbidity on individuals and healthcare systems^7^, multimorbidity-adjusted health risk assessment (HRA) tools^8–12^ have become fundamental instruments. Nevertheless, while the current approaches for HRA can certainly capture the burden of disease on individuals, they often fall short in envisaging disease progression and anticipating the onset of new comorbid conditions^13^. In contrast, the conceptualization of the diseasome^14^ sparked the appearance of a plethora of studies investigating the temporal patterns of disease concurrence, or disease trajectories^15,16^, yielding a better understanding of the time-dependent relationships among diseases and establishing a promising landscape to identify disease-disease causal relationships. Notably, these relationships are not arbitrary and frequently align with shared risk factors and/or underlying pathophysiological mechanisms^17–19^. Nevertheless, the traditional method of detecting disease co-occurrence has been deemed flawed as it may inadvertently generate false correlations between diseases, arising indirectly through multiple pairwise comparisons, exponentially increasing as the number of diseases examined escalates^20^. In this regard, sparse Bayesian Direct Multimorbidity Maps (BDMMs)^20,21^ showed to be a promising solution by filtering indirect disease associations.

The current observational retrospective multicentric cohort study employed BDMMs to investigate temporal disease maps among MDD and highly prevalent disease conditions^22^ in the context of the ERAPERMED EU project TRAJECTOME^23^. The study combined the results of the temporal disease maps identified in TRAJECTOME and the disability weights (DW)^24^ documented in the 2019 revision of the Global Burden of Diseases study (GBD), to develop and validate a Multimorbidity Adjusted Disability Score (MADS). The DW represents the degree of health loss caused by a specific disease. Our objective was twofold: 1) Identifying patients with different profiles of risk and assessing the disease burden of MDD and its comorbidities on individuals and health systems; and 2) Estimating the risk of morbidity progression and the onset of MDD comorbid conditions.

The development and evaluation of MADS involved the following steps:

**Step 1** – Computing age-dependent disease-disease probabilities of relevance (PR) using the BDMM method in four age intervals (0-20, 0-40, 0-60, and 0-70 years). This analysis resulted in an inhomogeneous dynamic Bayesian network that determined the PR for MDD against the most prevalent co-occurring diseases in the three European cohorts considered in TRAJECTOME, namely: The Catalan Health Surveillance System (CHSS)^25^, the UK Biobank (UKB)^26^, and The Finnish National Institute for Health and Welfare cohort (THL)^27^. THL cohort amalgamates information from Finrisk^28^ 1992, 1997, 2002, 2007, 2012, Finhealth^29^ 2017 and Health^30^ 2000/2011 studies.

**Step 2** – Combining the PR of every disease condition assessed in the study with their correspondent DW, extracted from the GBD 2019 study, we estimated the morbidity burden caused by MDD and its comorbid conditions. MADS was computed following a multiplicative combination of PR and DW of all the disease conditions present in an individual.

**Step 3** – Using MADS to stratify patients into different risk levels corresponding to different percentiles of the population-based risk pyramid of each patient cohort.

**Step 4** – Finally, the correspondence between the MADS risk strata and health outcomes were analysed through a cross-sectional analysis of utilisation of healthcare resources, mortality, pharmacological burden, and healthcare expenditure, and a longitudinal analysis of disease prevalence and incidence of new disease onsets. The results were validated through a multicentric replication of the findings in the three study cohorts, including 1,041,014 individuals.

## RESULTS

### Sociodemographic characteristics of the study cohorts

One of the first results is the characterisation of the three study cohorts and compared the sociodemographic attributes of their MADS risk groups (**Table 1**). All the individuals were classified into distinct risk strata based on quantiles of MADS distribution within the source population, resulting in the formation of the subsequent risk pyramid: Very low risk tier (≤ P_50_); Low risk tier (P_50_-P_80_]; Moderate risk tier (P_80_-P_90_]; High risk tier (P_90_-P_95_]; Very high risk tier (> P_99_).

To comprehend the inherent sociodemographic disparities across the cohorts under study, it is imperative to underscore the fundamental distinctions in their composition. Specifically, the THL and UKB cohorts predominantly consist of data derived from biobanks with a specific focus on the middle-aged and elderly population. In contrast, the CHSS cohort represents a population-based sample that encompasses all the population spectrum.

It is worth noting that a common pattern is observed among all the cohorts in the age distribution of the citizens at-risk. Although MADS is an additive morbidity grouper, it is not monotonically increasing with age. Remarkably, a notable proportion of high-risk cases were observed within the age range of 40 to 60 years, when depression typically manifests for the first time on average.

A divergence in the sex distribution across the risk strata is observable and especially noticeable in CHSS and UKB cohorts where the morbidity burden associated with depression and its related diseases is amplified women (P<.001). Likewise, the disability caused by depression and its comorbidities is larger in families with fewer economic resources (P<.001). Overall, the prevalence of MDD is greater in UKB than in the other cohorts. However, upon analysing the allocation of the population afflicted with depression in the risk pyramid, a total of 22,238 individuals (57.79% of those diagnosed with MDD) are categorized in the “high” and “very high” risk tiers in the CHSS cohort, whereas the number of individuals diagnosed with MDD that are allocated at the tip of the risk pyramid is 920 (40.22%) in THL and 23,409 (43.78%) in UKB.

**Table 1.**
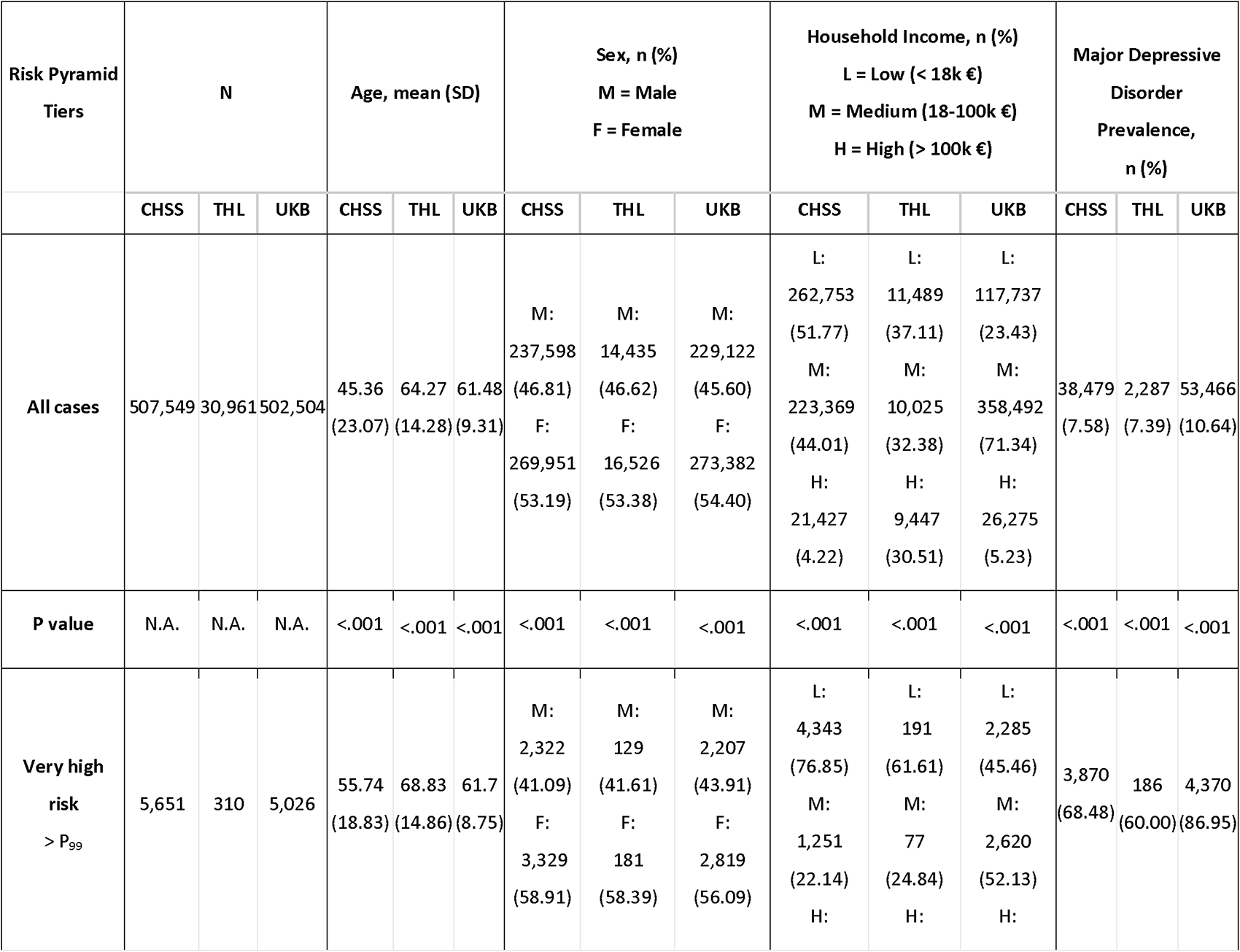
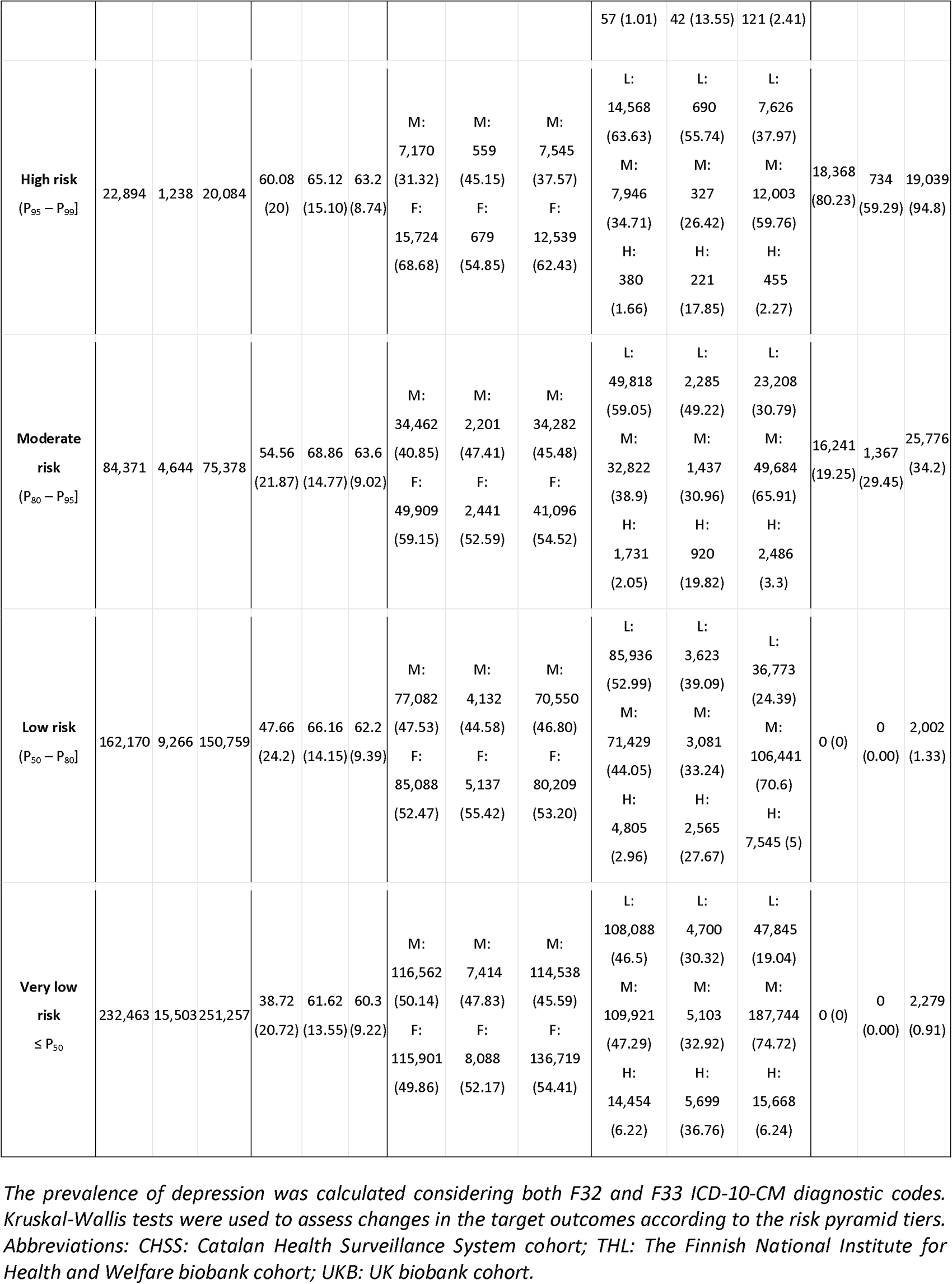
Demographic characteristics of each stratum of the MADS risk pyramid in the three study cohorts: CHSS^25^, UKB^26^ and THL^27^.

### Assessment of the MADS risk groups

#### Assessment of the PRs

Analysing the relationship between MDD and the morbidities assessed in the study is essential to interpret the MADS risk strata. This analysis revealed various relevant connections between MDD and the diseases investigated, encompassing both acute and chronic conditions, with the latter being particularly noteworthy due to their non-transient nature. Notably, the cluster of mental and behavioural disorders showed the highest average PRs in depression, but relevant associations also emerge among MDD and specific chronic somatic diseases affecting multiple organic systems (**Figure 1**).

**Figure 1.**
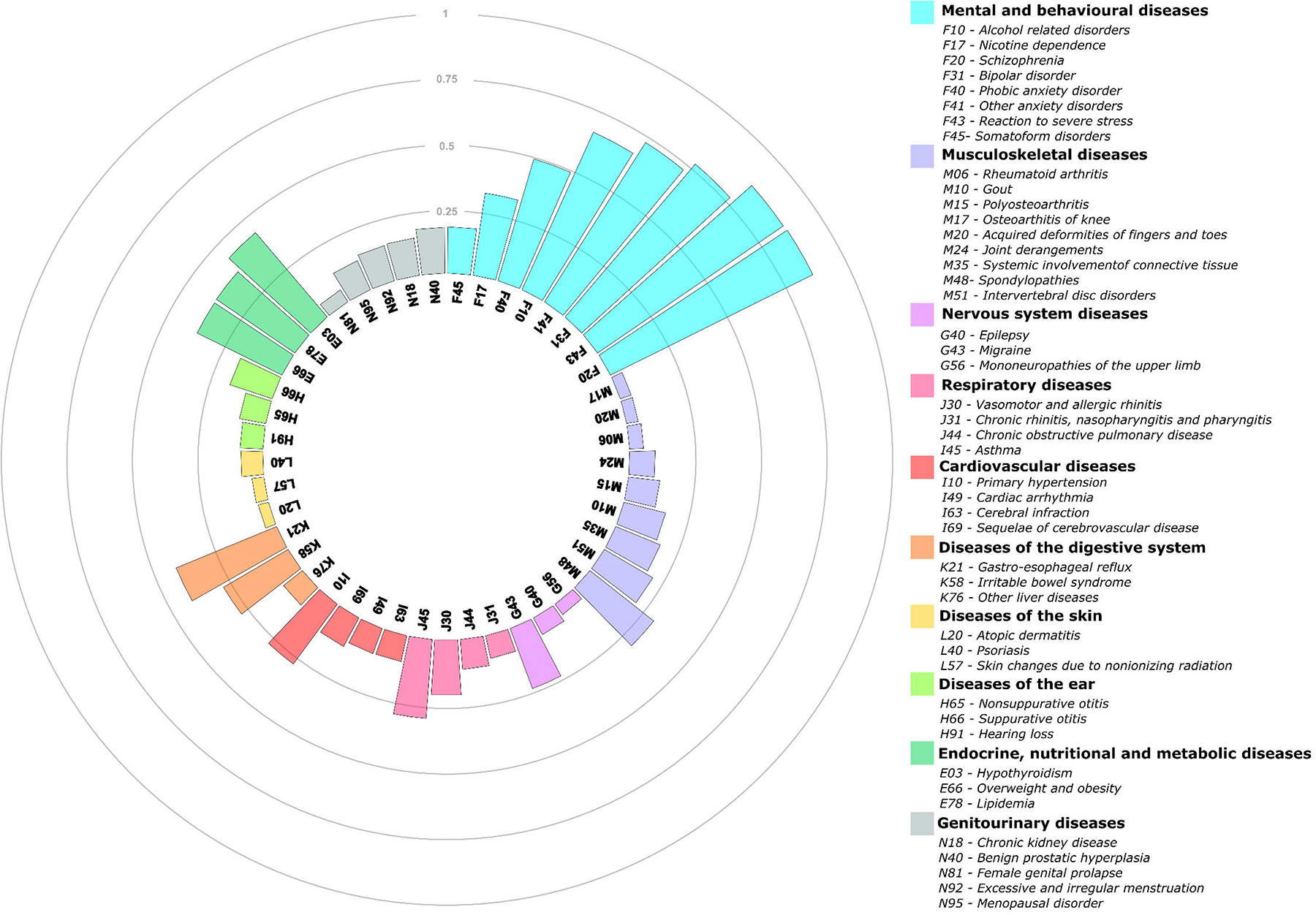
Average probabilities of relevance between Major Depressive Disorder and 45 chronic conditions utilized to compute MADS.

#### Utilisation of healthcare resources

The evaluation of the impact of MADS risk groups on healthcare systems was conducted by investigating the correlation between the MADS risk categories and the utilization of health resources over the 12-month period following the MADS assessment within the CHSS cohort (**Table 2**). The results illustrate a significant and gradual pattern of increased healthcare utilization as individuals progress from lower MADS risk tiers to higher risk tiers, reflecting an escalation in healthcare needs and requirements. Overall, patients with higher MADS scores exhibit a greater likelihood of experiencing morbidity-related adverse events, which subsequently leads to recurrent interactions with healthcare systems across multiple levels. These interactions include higher frequencies of primary care visits (P<.001), specialized outpatient visits (P<.001), emergency room visits (P<.001), hospital admissions (P<.001) and ambulatory visits in mental health centres (P<.001) as well as an increased pharmacological burden (P<.001).

**Table 2.** Utilization of healthcare resources over 12 months in each stratum of the MADS risk pyramid for the CHSS cohort.

| Risk Pyramid Tiers | Primary Care visits (per person) | Specialized Outpatient visits (per person) | Emergency Room visits (visits/100 inhabitants) | Hospital admissions (admissions/100 inhabitants) | Mental Health visits (visits/100 inhabitants) | Number of prescriptions (per person) |
| --- | --- | --- | --- | --- | --- | --- |
| **P value** | <.001 | <.001 | <.001 | <.001 | <.001 | <.001 |
| **Very high risk**<br>> $P_{99}$ | 12.50 | 3.07 | 135.00 | 28.50 | 554.00 | 8.02 |
| **High risk**<br>($P_{95} - P_{99}$] | 11.90 | 2.56 | 87.20 | 20.60 | 136.00 | 7.48 |
| **Moderate risk**<br>($P_{80} - P_{95}$] | 9.03 | 1.82 | 61.90 | 14.50 | 44.20 | 5.11 |
| **Low risk**<br>($P_{50} - P_{80}$] | 6.21 | 1.21 | 42.40 | 8.87 | 15.10 | 3.20 |
| **Very low risk**<br>$\leq P_{50}$ | 2.96 | 0.50 | 23.40 | 3.25 | 5.96 | 1.07 |

Kruskal-Wallis tests were used to assess changes in the target outcomes according to the risk pyramid tiers.

#### Mortality and healthcare expenditure

Moreover, we performed a cross-sectional analysis investigating mortality rates and the healthcare expenditure within the 12 months following the MADS assessment, expressed as the average healthcare expenditure per capita and differentiating among pharmaceutical and non-pharmaceutical costs, within the CHSS and THL cohorts (**Table 3**). The reported mortality rates (P<.001) were 5 to 20 times higher in the high-risk strata than in low-risk individuals. Likewise, the average healthcare expenditure per person, comprising both pharmacological (P<.001) and non-pharmacological spending (P<.001), was significantly higher for the individuals allocated at the tip of the risk pyramid than for those allocated at the bottom of the pyramid.

**Table 3:** Mortality rates and pharmacological and non-pharmacological healthcare expenditure in €, over 12 months, in each stratum of the MADS risk pyramid in CHSS^25^ and THL^27^.

| Risk Pyramid Tiers | Mortality<br>(cases/1k inhabitants) |  | Pharmacological<br>expenditure in €<br>(per person) |  | Hospitalization<br>expenditure in €<br>(per person) |  | Total expenditure in €<br>(per person) |
| --- | --- | --- | --- | --- | --- | --- | --- |
|  | CHSS | THL | CHSS | THL | CHSS | THL | CHSS |
| **P value** | <.001 | <.001 | <.001 | <.001 | <.001 | <.001 | <.001 |
| **Very high risk**<br>> P<sub>99</sub> | 46.2 | 36.0 | 1,214 | 966 | 539 | 270 | 12,517 |
| **High risk**<br>(P<sub>95</sub> – P<sub>99</sub>] | 41.5 | 33.7 | 772 | 1,131 | 383 | 340 | 8,404 |
| **Moderate risk**<br>(P<sub>80</sub> – P<sub>95</sub>] | 25.5 | 32.2 | 485 | 1,077 | 270 | 254 | 5,209 |
| **Low risk**<br>(P<sub>50</sub> – P<sub>80</sub>] | 11.5 | 14.8 | 292 | 810 | 165 | 185 | 3,075 |
| **Very low risk**<br>≤ P<sub>50</sub> | 2.57 | 7.3 | 99 | 363 | 60 | 123 | 1,192 |

Kruskal-Wallis tests and Fisher exact tests were used to assess changes in the target outcomes according to the risk pyramid tiers. Abbreviations: CHSS: Catalan Health Surveillance System cohort; THL: The Finnish National Institute for Health and Welfare biobank cohort.

#### Pharmacological burden

The study also examined the pharmacological burden on individuals after a 12-month period following the MADS assessment (**Table 4**). The findings indicate a significant positive association between the various risk strata and heightened pharmaceutical utilization, which is consistently observed across all cohorts. Specifically, individuals allocated at the top of the risk pyramid demonstrate significantly higher utilization of antidepressants (P<.001), antipsychotics (P<.001), anxiolytics (P<.001), and sedatives (P<.001) compared to those in lower risk categories, leading to a surge in the cost of medication.

**Table 4:** Prescription of depression related pharmacological treatments over 12 months in each stratum of the MADS risk pyramid in CHSS^25^, UKB^26^ and THL^27^.

| Risk Pyramid Tiers | Antipsychotic (N05A)<br>(per person) |  |  | Anxiolytic (N05B)<br>(per person) |  |  | Hypnotics and sedatives (N05C)<br>(per person) |  |  | Antidepressant (N06A)<br>(per person) |  |  |
| --- | --- | --- | --- | --- | --- | --- | --- | --- | --- | --- | --- | --- |
|  | CHSS | THL | UKB | CHSS | THL | UKB | CHSS | THL | UKB | CHSS | THL | UKB |
| **P value** | <.001 | <.001 | <.001 | <.001 | <.001 | <.001 | <.001 | <.001 | <.001 | <.001 | <.001 | <.001 |
| **Very high risk**<br>> P<sub>99</sub> | 0.75 | 0.60 | 0.33 | 0.47 | 0.21 | 0.27 | 0.15 | 0.14 | 0.24 | 0.79 | 0.43 | 0.80 |
| **High risk**<br>(P<sub>95</sub> – P<sub>99</sub>] | 0.20 | 0.27 | 0.18 | 0.46 | 0.19 | 0.20 | 0.10 | 0.12 | 0.19 | 0.66 | 0.41 | 0.71 |
| **Moderate risk**<br>(P<sub>80</sub> – P<sub>95</sub>] | 0.07 | 0.08 | 0.15 | 0.28 | 0.08 | 0.16 | 0.05 | 0.10 | 0.18 | 0.27 | 0.27 | 0.54 |
| **Low risk**<br>(P<sub>50</sub> – P<sub>80</sub>] | 0.03 | 0.03 | 0.13 | 0.14 | 0.04 | 0.12 | 0.02 | 0.07 | 0.13 | 0.08 | 0.11 | 0.36 |
| **Very low risk**<br>≤ P<sub>50</sub> | 0.01 | 0.01 | 0.11 | 0.04 | 0.02 | 0.09 | 0.01 | 0.04 | 0.10 | 0.02 | 0.06 | 0.26 |

For recurrently dispensed medication only the first prescription was considered in the analysis. Kruskal-Wallis tests were used to assess changes in the target outcomes according to the risk pyramid tiers. Abbreviations: CHSS: Catalan Health Surveillance System cohort; THL: The Finnish National Institute for Health and Welfare biobank cohort; UKB: UK biobank cohort.

To evaluate the influence of age and sex on the outcomes examined in this section, we replicated all the previously presented results, categorizing the outcomes by sex and age and reported them in the **Supplementary material – Appendix 1**. The results suggest that the morbidity burden in individuals might be a primary driver influencing the occurrence of adverse health events and the heightened utilization of healthcare resources.

#### Multimorbidity progression

We performed a longitudinal analysis in the CHSS cohort for investigating the prevalence and incidence of new MDD-associated diagnoses and its highly relevant comorbid conditions in 5-year intervals after MADS assessment for depression throughout the patients’ lifespan (**Figure 2**), allowing for a comprehensive examination of disease patterns over time. **Figure 2** only displays a representative selection of the results, the plots for all the disease conditions analysed in this study, as well as the results found in THL and UKB cohorts, are reported in the **Supplementary material – Appendix 2**.

In general, both MDD (**Figure 2** – **Panel A**), and the comorbid conditions investigated in this study exhibit a positive correlation between the MADS risk tiers and the current prevalence and incidence of new disease onsets within a subsequent 5-year interval. Notably the prevalence of the studied diseases is significantly higher than the population average in the high-risk groups. Distinct patterns are discernible for certain disorders. For instance, conditions characterized by minimal disability, such as, gastro-oesophageal reflux (**Figure 2** – **Panel D**), insomnia, back pain, and overweight, exhibit consistent upward trends, with both incidence and prevalence steadily increasing throughout the lifespan. In contrast, more disabling diseases like schizophrenia (**Figure 2** – **Panel B**), bipolar disorder, and alcohol abuse (**Figure 2** – **Panel C**), which precipitate rapid health deterioration and reduce life expectancy, attain peak prevalence and incidence levels during middle-aged adulthood, followed by a decline in later stages of life. These findings suggest a premature mortality among individuals afflicted by such conditions.

It is worth noting that in certain age intervals, the incidence of new MDD-related disease onsets is higher in the high-risk group compared to the very high-risk group. This phenomenon is explained due to the method employed to calculate the MADS, as the high-risk group primarily comprises individuals with highly disabling diseases (high DW) that are closely associated with depression (high PR), such as schizophrenia or bipolar disorder. It is also noteworthy that the 5-years incidence of certain disorders is very low. Considering that the high-risk and very high-risk groups represent only 4% and 1% of the sample size respectively, the effects of the variance are especially noticeable, leading to the observable pronounced sawtooth patterns on the chart.

**Figure 2.**
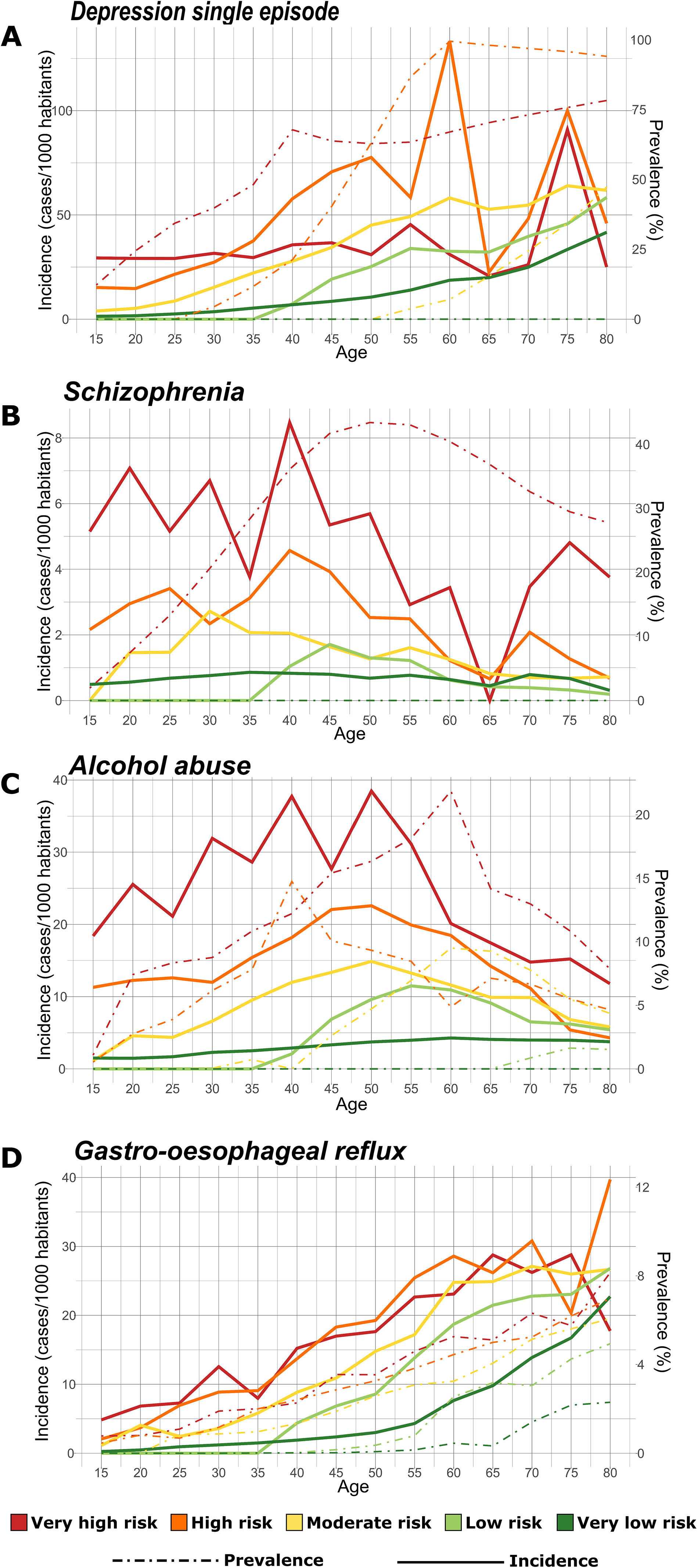
Longitudinal analysis of disease prevalence and incidence of new disease onsets. In *CHSS* cohort in four target disease conditions: Panel A) MDD single episode (ICD-10-CM: F32); Panel B) schizophrenia (ICD-10-CM: F20); Panel C) mental disorders related to alcohol abuse (ICD-10-CM: F10); Panel D) gastro-oesophageal reflux (ICD-10-CM: K21). Disease incidence is assessed in a 5-years interval, plotted in the left y-axis and represented with solid lines. Disease prevalence is plotted in the right y-axis and represented with dashed lines. The line colours correspond to the MADS risk pyramid tiers: red: very high-risk group; orange: high-risk group; yellow: medium-risk group; light green: low-risk group; green: very low-risk group.

## DISCUSSION

### Main findings

MADS seems to provide a unique and more comprehensive understanding of the complex nature of depression- related multimorbidity. This approach recognizes that individuals with depression often experience a range of comorbid conditions that may manifest and evolve differently over time. By capturing this dynamic aspect, MADS offers a nuanced assessment that goes beyond a mere checklist of discrete disorders. The novelty of the MADS approach lies in its capability to serve as the first morbidity grouper that incorporates information pertaining to disease trajectories, while improving the filtering of indirect disease associations using BDMMs.

In the current investigation, we have unearthed robust correlations between the MADS risk strata and the extent of deleterious impact caused by MDD and its comorbid conditions. Such associations indicate the presence of specific health risks and an escalated utilization of healthcare resources. Furthermore, a positive association has emerged between the levels of pharmacological and non-pharmacological healthcare expenditures and the different tiers of MADS risk. Also, the analysis has revealed an augmented risk of disease progression within the high-risk groups (high and very high-risk), as indicated by a heightened incidence of new-onset depression-related illnesses within a 12-month period after MADS assessment. Similarly, mortality rates have exhibited elevated values in these high-risk groups.

The findings presented in this study are underpinned by the complementary studies conducted within the TRAJECTOME project^22^ that have established a better understanding of the complex multimorbidity landscape associated with MDD across an individual’s lifespan, encompassing both modifiable and genetic risk factors.

### Potential impact in personalized medicine

By assessing whether MADS is appropriate for stratification of depression-related multimorbidity, we attempted to confirm its potential for contributing to precision medicine of patients with MDD who are in their early stages of MDD development^31^. Despite all advances toward adopting a personalised approach in mental health services, and more specifically in the treatment of depressive disorders, several challenges still limit its clinical utility^32^. The data-driven categorization of MADS in clinical practice may help facilitate further screenings and referrals of patients in a cost-effective manner.

The results reported in this study not only reaffirm the well-established link between multimorbidity and adverse outcomes such as a decline in functional status, compromised quality of life, and increased mortality rates^33^ but also shed light on the significant burden imposed on individuals and healthcare systems. The strain on resource allocation and overall healthcare spending is a pressing concern that necessitates effective strategies for addressing and managing multimorbidity^34^.

In this context, the assessment of individual health risks and patient stratification emerges as crucial approaches that enable the implementation of predictive and preventive measures in healthcare. By identifying individuals at higher risk and tailoring interventions accordingly, healthcare providers can proactively intervene, potentially averting or mitigating the progression of diseases and optimizing patient outcomes. These strategies not only yield immediate value in terms of improved patient care but also lay the foundation for the broader adoption of integrated care and precision medicine, particularly in the management of chronic conditions^35^.

Moreover, the findings of this study highlight the potential of preventive strategies targeted at mental disorders, including substance abuse disorders, depressive disorders, and schizophrenia, to reduce the incidence of negative clinical outcomes in somatic health conditions. These important implications for clinical practice call for a comprehensive and interdisciplinary approach that bridges the gap between psychiatric and somatic medicine. By developing cross-specialty preventive strategies, healthcare professionals can provide more holistic and effective care for individuals with complex health needs, ensuring that both their mental and physical health are adequately addressed^4^.

### Further developments

The methodological approach used to develop MADS has proven to be effective in measuring the impact of an index disease and its principal comorbidities both in individuals and healthcare systems.

Furthermore, it is expected that MADS approach might be used in other well-established clusters of non-communicable diseases. By leveraging this targeted approach, MADS can be adapted to other disease clusters with shared characteristics, enabling a more precise assessment of disease burden and comorbidity patterns. Since MADS was constructed using only the calculations derived from the BDMM analysis of 86 disease conditions previously screened from a large set of disease conditions, MADS focuses only on the health problems directly related to the index disease (i.e., MDD) and ignores the surrounding effects of the uncorrelated comorbid conditions.

Finally, MADS, or derived scores, can be integrated as part of holistic strategies for subject-specific risk assessment that combines information on various patient’s determinants of health, also known as Multisource Clinical Predictive Modelling (MCPM). The integration of MADS with other pertinent risk factors and clinical information within MCPM approaches offers a comprehensive framework for conducting risk assessments and implementing personalized interventions in the clinical arena^36,37^. This statement was grounded on the hypothesis that the implementation of comprehensive strategies for subject-specific risk prediction and stratification, incorporating multiple sources of covariates influencing patients’ health, could enhance the accuracy of predictions and facilitate informed clinical decision-making by providing reliable estimates of individual prognosis^38^. This strategy is expected to support the implementation and long-term use of preventive measures for managing chronic patients, with the goal of delaying or preventing their progression to the highest risk level on the stratification pyramid^39^.

The current study also provided good prospects for utilization of the disease trajectories to enhance the performance of existing state-of-the-art morbidity groupers, such as the Adjusted Morbidity Groups (AMG)^2,12^. AMG system is currently used in Catalonia (ES; 7M inhabitants), both for health policy and clinical purposes, and promoted for its transferability to other EU regions by the EU joint Action on implementation of digitally enabled integrated person-centred care (JADECARE)^40^. Unlike the current approach based on DW, the AMG score employs a disease-specific weighting derived from statistical analysis incorporating mortality and healthcare service utilization data. This approach would allow the creation of tools adjustable to the characteristics of each healthcare system, adapting to the impact of a particular disease condition into a specific region and improving the generalisation capacity of the tool.

### Limitations of the current approach

Despite meeting expectations and validating the hypothesis by which the study was conceived, the authors acknowledge a series of limitations leading to suboptimal results and limited potential for adaptation and generalization that should be undertaken to bring MADS, or an indicator derived from it to real world implementation.

In the current research, the use of estimations of mean DW^41^ to assess the burden of disease conditions has achieved desirable results, and is conceptually justified, but undoubtedly exhibits major limitations. In an ideal clinical scenario, each disease diagnosis indicated in the patient’s electronic medical record should be accompanied by the characterization of three key dimensions: i) severity of the diagnosis, ii) rate of disease progression, and iii) impact on disability. However, the degree of maturity for the characterization of the last two dimensions, disease progression and disability, is rather poor because of the complexities involved in their assessment. In other words, the authors are acknowledging the weakness associated with the current use of DW, but they are stressing the importance of incorporating such dimension, directly assessed on individual basis, in future evolutions of MADS.

A noteworthy aspect that should be acknowledged is that factors such as the advancement of diagnostic techniques, the digitization of medical records, and the modifications in disease taxonomy and classification over time, have contributed to a more exhaustive documentation of the disease states in the most recent health records. Consequently, this fact could lead to imprecisions estimating the disease onset ages in the older individuals.

## CONCLUSIONS

MADS showed to be a promising approach to estimate multimorbidity-adjusted risk of disease progression and measure MDD’s impact on individuals and healthcare systems, which could be tested in other diseases. The novelty of the MADS approach lies in its unique capability to incorporate disease trajectories, providing a comprehensive understanding of depression-related morbidity burden. In this regard, the BDMM method played a crucial role in isolating and identifying true direct disease associations. Nevertheless, clinical validation is imperative before considering the widespread adoption of MADS in routine clinical practice.

## METHODS

### Data sources

The study was conducted utilizing data from three public health cohorts, namely:

1. **The Catalan Health Surveillance System (CHSS)** – The main cohort used in MADS development was extracted from the *CHSS*. Operated by a single-public payer (CatSalut)^42^ since 2011, the *CHSS* gathers information across healthcare tiers on the utilization of public healthcare resources, pharmacological prescriptions, and patients’ basic demographic data, including registries of 7.5 million citizens from the entire region of Catalonia (ES). Nevertheless, for MADS development purposes we considered only registry data from the citizens resident in the entire Health District of Barcelona-Esquerra (AISBE) between 1^st^ of January of 2011 and 31^st^ of December of 2019 (n=654,913). To validate the results of MADS, we retrieved additional information from *CHSS* corresponding to the 12 months posterior to MADS assessment, from 1^st^ of January of 2020 to 31^st^ of December of 2020. It is to note that, all the deceased patients in addition to those who moved their residence outside of AISBE district between 2011 and 2019 were discarded from the validation analysis, the remaining subset of patients comprises 508,990 individuals.
2. **The United Kingdom Biobank (UKB)** – The UKB data considered in this study contained medical and phenotypic data from participants aged between 37-93 years. Recruitment was based on NHS patient registers and initial assessment visits were carried out between 3rd of March of 2006 and 1st of October of 2010 (n = 502,504). Analysed data included disease diagnosis and onset time, medication prescriptions, and socio-economic descriptors.
3. **The Finnish National Institute for Health and Welfare biobank (THL)** – THL cohort integrates information from Finrisk^28^ 1992, 1997, 2002, 2007, 2012, Finhealth^29^ 2017 and Health^30^ 2000/2011 studies. For the consensual clustering 41,092 participants were used from Finnish population surveys. After data cleaning, 30,961 participants remained from Finnish population surveys. These participants of age 20-100 were chosen at random from the Finnish population and represented different parts of Finland.

### Ethical approval

As a multicentric study, TRAJECTOME accessed multiple cohorts’ data, all subject to the legal regulations of their respective regions of origin and obtained the necessary approvals from the corresponding ethics committees.

For CHSS cohort, the Ethical Committee for Human Research at Hospital Clinic de Barcelona approved the core study of TRAJECTOME on the 24^th^ of March of 2021 (HCB/2020/1051) and subsequently approved the analysis for the generation and validation of MADS on the 25^th^ of July of 2022 (HCB/2022/0720).

UK Biobank received ethical approval from the National Research Ethics Service Committee Northwest– Haydock (ref. 11/NW/0382).

The THL cohort integrates information from the Finrisk databases: 1997 (Ethical committee of National Public Health Institute. Statement 38/96. 30.10.1996), 2002 (Helsinki University Hospital, Ethical committee of epidemiology and public health, Statement 87/2001. Reference 558/E3/2001. 19.12.2001), 2007 (Helsinki University Hospital, Coordinating ethics committee, Dnro HUS 229/EO/2006, 20.6.2006) and 2012 (Helsinki University Hospital, Coordinating ethics committee, Dnro HUS 162/13/03/11, 1.12.2011); the FinHealth 2017 (Helsinki University Hospital, Coordinating ethics committee, 37/13/03/00/2016 22.3.2016) and the Health 2000-2011 databases (Ethical committee of National Public Health Institute, 8/99/12. Helsinki University Hospital, Ethical committee of epidemiology and public health, 407/E3/2000. 31.05.2000 and 17.06.2011).

The ethics committees exempted the requirement to obtain informed consent for the analysis, and publication of retrospectively acquired and fully anonymized data in the context of this non-interventional study.

All the data was handled in compliance with the General Data Protection Regulation 2016/679, which safeguards data protection and privacy for all individuals in the European Union. The study was conducted in conformity with the Helsinki Declaration (Stronghold Version, Brazil, October 2013) and in accordance with the protocol and the relevant legal requirements (Biomedical Research Act 14/2007 of 3 July).

### Building the Multimorbidity Adjusted Disability Score

The development of MADS intertwined four development and evaluation steps (Figure 3): Step 1) Computing age-dependent disease-disease probabilities of relevance; Step 2) Extracting and aggregating the DW; Step 3) Generating the MADS risk pyramid; and Step 4). Evaluation of MADS risk strata.

**Figure 3.**
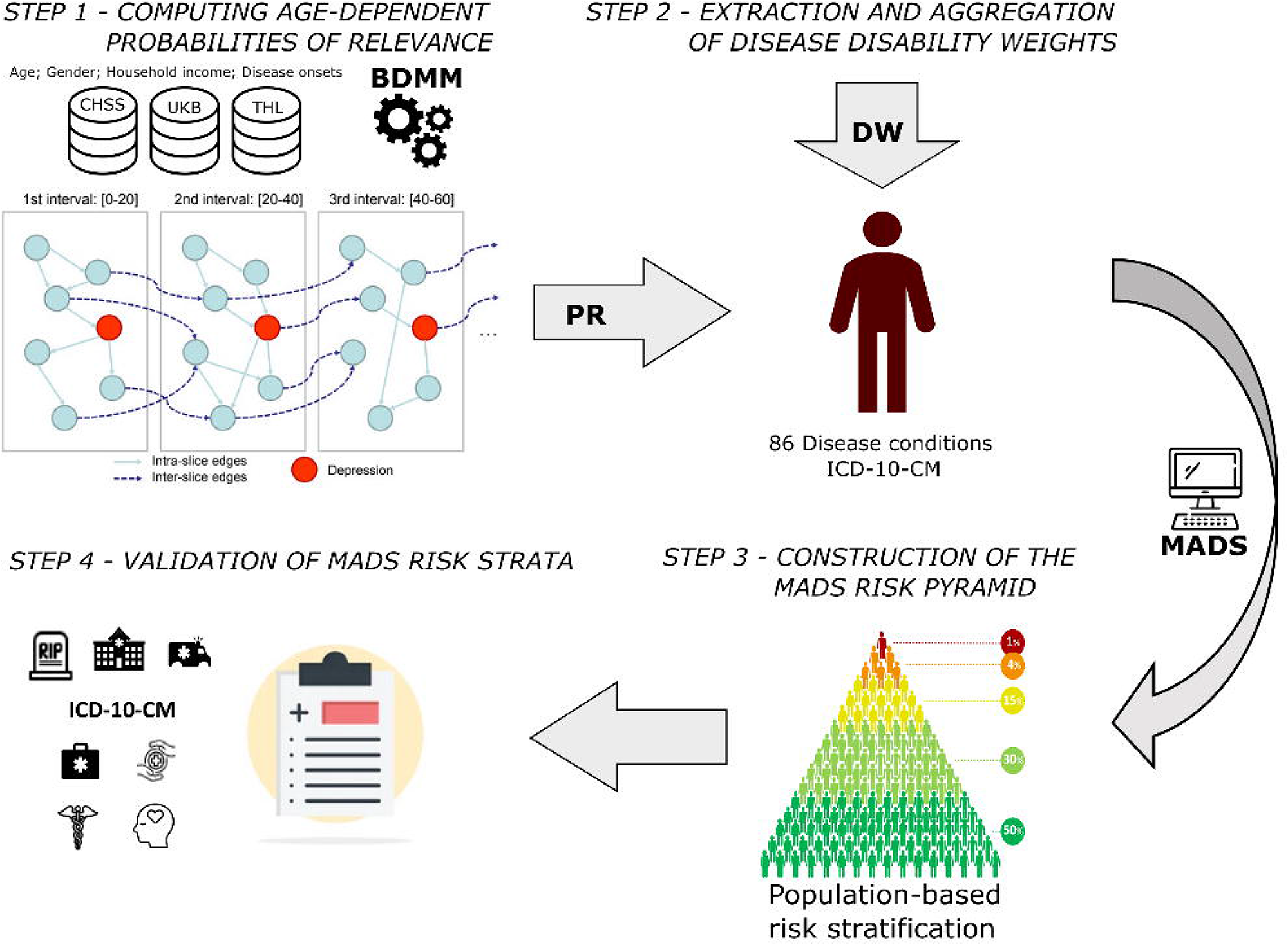
Workflow for building and validation of the MADS. BDMM stands for Bayesian Direct Multimorbidity Maps, PR for Probabilities of Relevance, and DW for Disability Weights.

#### STEP 1- COMPUTING AGE-DEPENDENT PROBABILITIES OF RELEVANCE

BDMMs were used to assess direct and indirect associations between MDD and a set of 86 potential comorbid conditions. The set of 86 disease conditions considered in the study had a prevalence greater than 1% in all the study cohorts. The list of diseases and their associated ICD-10-CM^3^ codes are displayed in the **Supplementary material – Appendix 3**.

This step considered information on: 1) **Disease diagnosis:** Disease conditions were catalogued using the first three characters of ICD-10-CM codes; 2) **Age at disease onset time:** The age at disease onset corresponds to the first diagnosis in a lifetime for each ICD-10-CM code; 3) **Sex**; and, 4) **Socio-economic status**: annual average total household income (before tax with co-payment exemption) as a categorical variable with 3 categories: a) Less than 18,000; b) 18,000 to 100,000; c) Greater than 100,000. Thresholds are given in EUR.

BDMM analysis resulted in an inhomogeneous dynamic Bayesian network, which was utilised to compute temporal PR, ranging from 0 (no association) to 1 (strong association), for MDD in conjunction with sex, socio- economic status, and the set of 86 predetermined consensual diseases^22^. To construct the trajectories, the PR was calculated in four different age ranges: 0-20, 0-40, 0-60, and 0-70 years of age. The PR calculated and utilized for MADS computation are reported in the **Supplementary material – Appendix 4**. Further details regarding the core analysis conducted in TRAJECTOME can be found in ^22^.

#### STEP 2 - EXTRACTION AND AGGREGATION OF DISEASE DISABILITY WEIGHTS (DW)

MADS was developed by weighting the DWs of single diseases according to their estimated PR against MDD. DWs indicate the degree of health loss, based on several health outcomes, and are used as indicators of the total of disability caused by a certain health condition or disease. Often, the DWs present specific disability scores tailored to the severity of the disease. The disease categories, their severity distribution and their associated DWs utilised in this study were extracted from the GBD studies of 2019 and reported in the **Supplementary material – Appendix 3**.

DWs were extracted and aggregated as follows: 1) We considered only the DW of MDD and the set of 86 disease codes; 2) We considered the DW of all the chronic conditions diagnosed in patients’ lifetime, whereas, since the disability caused by acute illnesses is transitory, the DWs for the acute diseases diagnosed more than 12 months before the MADS assessment were arbitrary set to 0 (no disability); 3) Due to the unavailability of information on the severity of diagnoses, we determined the DWs of each disease condition by calculating the weighted mean of the DWs associated to the disease severity categories and their prevalence. In instances where the severity distribution was not available, we computed the arithmetic mean of the DWs of each severity category; 4) We finally weighted the DWs according to the PR of each disease condition with respect to MDD, the PR were adjusted according to age of disease onset, discretized in the following intervals 0-20, 20-40, 40-60, >60 years old.

Since the DWs do not account for multimorbidity in their estimates, the utilization of DW independently can cause inaccuracies in burden of disease estimations, particularly in ageing populations that include large proportions of persons with two or more disabling disease conditions^43^. Consequently, we combined the DW and the PR for all the disease conditions present in one individual following a multiplicative approach (**Eq. 1**)^44^, aggregating several DW in a single score that accounts for the overall disability caused by numerous concurrent chronic conditions, in which every comorbid disease increases the utility loss of a patient, though it is less than the sum of the utility loss of both diseases independently.

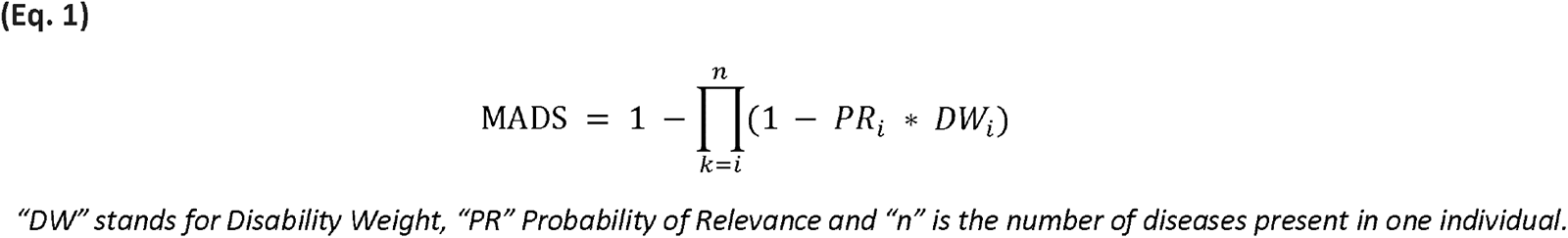

#### STEP 3 - CONSTRUCTION OF THE MADS RISK PYRAMID

Once calculated, MADS was utilised to stratify patients in different levels of risk according to the quantiles of its distribution in the source population, producing the following risk pyramid: 1) Very low risk (≤ P_50_); 2) Low risk (P_50_-P_80_]; 3) Moderate risk (P_80_-P_90_]; 4) High risk (P_90_-P_95_]; 5) Very high risk (> P_99_).

#### STEP 4 - EVALUATION OF MADS RISK STRATA

The validation of the results encompassed two interconnected analyses: 1) A cross-sectional validation of health outcomes; and 2) a longitudinal analysis of disease prevalence and incidence of new onsets.

#### CROSS-SECTIONAL VALIDATION OF HEALTH OUTCOMES AND USE OF HEALTHCARE RESOURCES

To validate the results of MADS, we conducted a cross-sectional analysis of clinical outcomes within the 12 months following the MADS assessment. The burden of MDD and its comorbidities on patients and healthcare providers, corresponding to each risk group of the MADS risk pyramid, was assessed using the following features (the parameters evaluated in each cohort may vary depending on the availability of the requested information in the source databases):

1. **Prescriptions of psycholeptic and psychoanaleptic drugs** (Information available in all the databases) The prescribed medication was catalogued using the first 4 characters from ATC^45^ codes, resulting in the following categories: *Antipsychotics (N05A), Anxiolytics (N05B), Hypnotics and sedatives (N05C) and Antidepressants (N06A)*.
2. **Cost of the pharmacological prescriptions in €** (Information available only in *CHSS* and *THL*).
3. **Mortality rates** (Information available only in *CHSS* and *THL*).
4. **Contacts and encounters with healthcare professionals** (Information available only in *CHSS*) Encompassing: i) primary care visits; ii) specialised care outpatient visits; iii) ambulatory visits in mental health centres; iv) emergency room visits; v) planned and unplanned hospital admissions; and vi) admissions in mental health centres.
5. ***Total healthcare expenditure*** (Information available only in *CHSS*) Including: i) direct healthcare delivery costs; ii) pharmacological costs; and iii) other billable healthcare costs, such as non-urgent medical transportation, ambulatory rehabilitation, domiciliary oxygen therapy, and dialysis.

We assessed the effect of sex and age replicating the analyses disaggregated by sex and age. The age ranges were discretized in the following categories: 0-20, 20-40, 40-60, >60 years.

#### LONGITUDINAL ANALYSIS OF DISEASE PREVALENCE AND INCIDENCE OF NEW ONSETS

To address the age-dependency of disease onsets we performed a longitudinal analysis of the prevalence of a target disease and the incidence of new diagnostics within the 5 years following the MADS assessment.

We iteratively computed MADS in five-year intervals throughout the patients’ life. Within each interval, the population was stratified based on the MADS distribution. Subsequently, within each risk tier, the prevalence of the target disease and the incidence of new disease onset over the subsequent five years were calculated. Only individuals with complete information for the next interval at each timepoint of the analysis were included.

In the analysis we considered only the disease conditions with a PR against MDD ≥ 0.80 in at least one of the four age intervals assessed, namely: 0-20, 0-40, 0-60 and 0-70. Resulting in the following set of mental diseases: *MDD (F32-F33), schizophrenia (F20), bipolar disorder (F31), anxiety related disorders (F40-F41), stress related disorders (F43), mental disorders related to alcohol abuse (F10), insomnia (G47)*. And the following somatic diseases: *dorsalgia (M54), soft tissue disorders not classified elsewhere (M79), irritable bowel syndrome (K58), overweight and obesity (E66) and gastro-oesophageal reflux (K21)*.

### Statistical analysis

Mortality rates were summarised as cases per 1,000 inhabitants, whereas numeric health outcomes variables were described by the average number of cases per person, per 100 inhabitants or per 1,000 inhabitants according to their prevalence. Average healthcare expenditures are reported in € per person. To evaluate changes in the target outcomes across the risk pyramid tiers, Kruskal-Wallis tests and Fisher exact tests were employed accordingly. Statistical significance was determined by considering a P-value less than .05 in all analyses.

All the data analyses were performed using R^46^, version 4.1.1 (R Foundation for Statistical Computing, Vienna, Austria). The MADS algorithm was fully developed and tested in the *CHSS* database and transferred to the other sites through an R programming executable script.

The study is reported according to the STROBE^23^ guidelines for observational studies.

## Data Availability

Data for this study are not publicly available due to patient privacy concerns. The scripts used to compute MADS are available from the corresponding author upon reasonable request.

## ACKNOWLEDGMENTS

The initiative was supported by ERA PerMed program (TRAJECTOME project, ERAPERMED2019-108).

The Catalan cohort was extracted from the Catalan Health Surveillance System database, owned, and managed by the Catalan Health Service, with the earnest collaboration of the Digitalization for the Sustainability of the Healthcare (DS3) - IDIBELL group.

The study was funded by the Academy of Finland under the frame of ERA PerMed (TRAJECTOME project, ERAPERMED2019-108). We want to acknowledge the participants and investigators of the FinnGen study. The authors wish to acknowledge CSC – IT Center for Science, Finland, for computational resources.

This research has been conducted using the UK Biobank Resource under Application Number 1602. Linked health data Copyright © 2019, NHS England. Re-used with the permission of the UK Biobank. All rights reserved. This study was supported by the Hungarian National Research, Development, and Innovation Office 2019-2.1.7-ERA-NET-2020-00005 under the frame of ERA PerMed (ERAPERMED2019-108); the Hungarian National Research, Development, and Innovation Office (K 143391, K 139330 and PD 134449 grants); the Hungarian Brain Research Program 3.0 (NAP2022-I-4/2022); and the Ministry of Innovation and Technology of Hungary from the National Research, Development and Innovation Fund, under the TKP2021-EGA funding scheme (TKP2021-EGA-25 and TKP2021-EGA-02). Supported by the European Union project RRF-2.3.1-21-2022-00004 within the framework of the Artificial Intelligence National Laboratory

## Authors’ Contributions

All authors contributed to the writing of this paper and approved the final draft.

## Conflicts of Interest

None declared.

## ACRONYMS

AISBE: Health District of Barcelona-Esquerra
AMG: Adjusted Morbidity Groups
BDMM: Bayesian Direct Morbidity Maps
CHSS: Catalan Health Surveillance System
DW: Disability Weights
GBD: Global Burden of Disease
MADS: Multimorbidity Adjusted Disability Score
MCPM: Multisource Clinical Predictive Modelling
MDD: Major Depressive Disorder
PR: Probability of Relevance
THL: Finnish National Institute for Health and Welfare Biobank
UKB: United Kingdom Biobank

## Notes

### Competing Interest Statement

The authors have declared no competing interest.

### Author Declarations

As a multicentric study, TRAJECTOME accessed multiple cohorts data, all subject to the legal regulations of their respective regions of origin and obtained the necessary approvals from the corresponding ethics committees. For CHSS cohort, the Ethical Committee for Human Research at Hospital Clinic de Barcelona approved the core study of TRAJECTOME on the 24th of March of 2021 (HCB/2020/1051) and subsequently approved the analysis for the generation and validation of MADS on the 25th of July of 2022 (HCB/2022/0720). UK Biobank received ethical approval from the National Research Ethics Service Committee Northwest Haydock (ref. 11/NW/0382). The THL cohort integrates information from the Finrisk databases: 1997 (Ethical committee of National Public Health Institute. Statement 38/96. 30.10.1996), 2002 (Helsinki University Hospital, Ethical committee of epidemiology and public health, Statement 87/2001. Reference 558/E3/2001. 19.12.2001), 2007 (Helsinki University Hospital, Coordinating ethics committee, Dnro HUS 229/EO/2006, 20.6.2006) and 2012 (Helsinki University Hospital, Coordinating ethics committee, Dnro HUS 162/13/03/11, 1.12.2011), the FinHealth 2017 (Helsinki University Hospital, Coordinating ethics committee, 37/13/03/00/2016 22.3.2016) and the Health 2000/2011 databases (Ethical committee of National Public Health Institute, 8/99/12. Helsinki University Hospital, Ethical committee of epidemiology and public health, 407/E3/2000. 31.05.2000 and 17.06.2011). The ethics committees exempted the requirement to obtain informed consent for the analysis, and publication of retrospectively acquired and fully anonymized data in the context of this noninterventional study. All the data was handled in compliance with the General Data Protection Regulation 2016/679, which safeguards data protection and privacy for all individuals in the European Union. The study was conducted in conformity with the Helsinki Declaration (Stronghold Version, Brazil, October 2013) and in accordance with the protocol and the relevant legal requirements (Biomedical Research Act 14/2007 of 3 July).

