## Supplementary Material for "Multicentric validation of a Multimorbidity Adjusted Disability Score to stratify depression-related risks using temporal disease maps"

Rubèn González-Colom et al.

*(On-line supplementary material)*

Table of content

### Annex 1: Cross-sectional analysis of health outcomes and utilization of healthcare resources

#### 1.1 CHSS cohort

##### Table S1 – Analysis of average number of primary care visits person over the 12 months following MADS assessment itemized by age and sex categories in CHSS.

| MADS  Risk Pyramid Tiers | Primary Care visits  (per person)  Male 0-20 | Primary Care visits  (per person)  Male 20-40 | Primary Care visits  (per person)  Male 40-60 | Primary Care visits  (per person)  Male >60 | Primary Care visits  (per person)  Female 0-20 | Primary Care visits  (per person)  Female 20-40 | Primary Care visits  (per person)  Female 40-60 | Primary Care visits  (per person)  Female >60 |
| --- | --- | --- | --- | --- | --- | --- | --- | --- |
| Very high risk  > P_99_ | 5.38 | 7.4 | 10.49 | 14.61 | 7.76 | 11.18 | 12.83 | 15.58 |
| High risk  (P_95_ – P_99_] | 5.83 | 7.59 | 9.03 | 14.06 | 7.3 | 9.29 | 10.92 | 14.55 |
| Moderate risk  (P_80_ – P_95_] | 5.76 | 5.64 | 7.3 | 11.83 | 6.09 | 6.93 | 8.43 | 11.96 |
| Low risk  (P_50_ – P_80_] | 4.9 | 3.98 | 4.77 | 8.73 | 4.92 | 5.18 | 5.63 | 8.75 |
| Very low risk  ≤ P_50_ | 2.77 | 2.31 | 2.3 | 4.24 | 2.85 | 3.06 | 2.86 | 4.12 |

##### Table S2 - Analysis of average number of outpatient care visits person over the 12 months following MADS assessment itemized by age and sex categories in CHSS.

| MADS  Risk Pyramid Tiers | Specialized outpatient visits  (per person)  Male 0-20 | Specialized outpatient visits  (per person)  Male 20-40 | Specialized outpatient visits  (per person)  Male 40-60 | Specialized outpatient visits  (per person)  Male >60 | Specialized outpatient visits  (per person)  Female 0-20 | Specialized outpatient visits  (per person)  Female 20-40 | Specialized outpatient visits  (per person)  Female 40-60 | Specialized outpatient visits  (per person)  Female >60 |
| --- | --- | --- | --- | --- | --- | --- | --- | --- |
| Very high risk  > P_99_ | 1.45 | 2.45 | 3.07 | 3.28 | 3.59 | 2.38 | 3.75 | 3.05 |
| High risk  (P_95_ – P_99_] | 2.33 | 1.28 | 2.46 | 3.06 | 2.2 | 1.65 | 2.83 | 2.75 |
| Moderate risk  (P_80_ – P_95_] | 1.67 | 0.86 | 1.64 | 2.74 | 1.93 | 1.07 | 1.68 | 2.27 |
| Low risk  (P_50_ – P_80_] | 0.97 | 0.56 | 0.88 | 2.12 | 0.83 | 0.73 | 1.09 | 1.76 |
| Very low risk  ≤ P_50_ | 0.43 | 0.28 | 0.34 | 1 | 0.4 | 0.4 | 0.49 | 0.93 |

##### Table S3 - Analysis of average number of emergency room visits per 100 habitants over the 12 months following MADS assessment itemized by age and sex categories in CHSS.

| MADS  Risk Pyramid Tiers | Emergency room visits  (visits/100 hab.)  Male 0-20 | Emergency room visits  (visits/100 hab.)  Male 20-40 | Emergency room visits  (visits/100 hab.)  Male 40-60 | Emergency room visits  (visits/100 hab.)  Male >60 | Emergency room visits  (visits/100 hab.)  Female 0-20 | Emergency room visits  (visits/100 hab.)  Female 20-40 | Emergency room visits  (visits/100 hab.)  Female 40-60 | Emergency room visits  (visits/100 hab.)  Female >60 |
| --- | --- | --- | --- | --- | --- | --- | --- | --- |
| Very high risk  > P_99_ | 121.43 | 140.83 | 140.4 | 123.86 | 178.38 | 164.79 | 120.47 | 130.87 |
| High risk  (P_95_ – P_99_] | 76.6 | 94.32 | 77.36 | 91.05 | 128.99 | 102.61 | 70.86 | 91.01 |
| Moderate risk  (P_80_ – P_95_] | 57.12 | 58.73 | 54.11 | 71.87 | 66.76 | 68.81 | 49.56 | 66.8 |
| Low risk  (P_50_ – P_80_] | 52.4 | 39.45 | 31.84 | 46.94 | 48.42 | 52.13 | 32.99 | 42.74 |
| Very low risk  ≤ P_50_ | 28.46 | 25.1 | 17.91 | 22.15 | 25.51 | 31.22 | 17.71 | 19.15 |

##### Table S4 - Analysis of average number of hospital admissions per 100 habitants over the 12 months following MADS assessment itemized by age and sex categories in CHSS.

| MADS  Risk Pyramid Tiers | Hospital admissions  (admissions/100 hab.)  Male 0-20 | Hospital admissions  (admissions/100 hab.)  Male 20-40 | Hospital admissions  (admissions/100 hab.)  Male 40-60 | Hospital admissions  (admissions/100 hab.)  Male >60 | Hospital admissions  (admissions/100 hab.)  Female 0-20 | Hospital admissions  (admissions/100 hab.)  Female 20-40 | Hospital admissions  (admissions/100 hab.)  Female 40-60 | Hospital admissions  (admissions/100 hab.)  Female >60 |
| --- | --- | --- | --- | --- | --- | --- | --- | --- |
| Very high risk  > P_99_ | 14.29 | 23.55 | 25.53 | 40.97 | 27.03 | 23.48 | 21.53 | 34.63 |
| High risk  (P_95_ – P_99_] | 7.23 | 8.52 | 16.5 | 35.99 | 15.65 | 9.97 | 12.17 | 26.76 |
| Moderate risk  (P_80_ – P_95_] | 6.16 | 5.07 | 10.49 | 30.61 | 6.62 | 7.66 | 8.35 | 20.8 |
| Low risk  (P_50_ – P_80_] | 3.71 | 3.52 | 6.04 | 20.12 | 2.78 | 6.24 | 5.58 | 14.26 |
| Very low risk  ≤ P_50_ | 1.81 | 1.8 | 2.58 | 8.42 | 1.3 | 3.74 | 2.68 | 5.66 |

##### Table S5 - Analysis of average number of mental health visits per 100 habitants over the 12 months following MADS assessment itemized by age and sex categories in CHSS.

| MADS  Risk Pyramid Tiers | Mental Health visits  (visits/100 hab.)  Male 0-20 | Mental Health visits  (visits/100 hab.)  Male 20-40 | Mental Health visits  (visits/100 hab.)  Male 40-60 | Mental Health visits  (visits/100 hab.)  Male >60 | Mental Health visits  (visits/100 hab.)  Female 0-20 | Mental Health visits  (visits/100 hab.)  Female 20-40 | Mental Health visits  (visits/100 hab.)  Female 40-60 | Mental Health visits  (visits/100 hab.)  Female >60 |
| --- | --- | --- | --- | --- | --- | --- | --- | --- |
| Very high risk  > P_99_ | 1430.95 | 923.85 | 706.77 | 318.9 | 818.92 | 584.15 | 654.48 | 278.71 |
| High risk  (P_95_ – P_99_] | 522.98 | 137.33 | 266.19 | 51.6 | 520.58 | 138.01 | 206.16 | 54.03 |
| Moderate risk  (P_80_ – P_95_] | 194.13 | 47.65 | 54.2 | 12.57 | 220.36 | 39.88 | 43.19 | 18.47 |
| Low risk  (P_50_ – P_80_] | 54 | 12.67 | 10.4 | 4.55 | 37.33 | 14.14 | 10.24 | 5.73 |
| Very low risk  ≤ P_50_ | 27.2 | 2.52 | 1.71 | 0.77 | 14.67 | 3.12 | 1.4 | 1.17 |

##### Table S6 - Analysis of average number of different pharmacological prescriptions per person over the 12 months following MADS assessment itemized by age and sex categories in CHSS.

| MADS  Risk Pyramid Tiers | Number of prescriptions  (per person)  Male 0-20 | Number of prescriptions  (per person)  Male 20-40 | Number of prescriptions  (per person)  Male 40-60 | Number of prescriptions  (per person)  Male >60 | Number of prescriptions  (per person)  Female 0-20 | Number of prescriptions  (per person)  Female 20-40 | Number of prescriptions  (per person)  Female 40-60 | Number of prescriptions  (per person)  Female >60 |
| --- | --- | --- | --- | --- | --- | --- | --- | --- |
| Very high risk  > P_99_ | 3.19 | 3.99 | 6.87 | 9.85 | 4.85 | 5.61 | 8.32 | 10.79 |
| High risk  (P_95_ – P_99_] | 1.89 | 2.73 | 5.31 | 10.03 | 3.28 | 3.77 | 6.2 | 10.22 |
| Moderate risk  (P_80_ – P_95_] | 1.59 | 1.82 | 3.58 | 8.45 | 1.98 | 2.5 | 4.02 | 8.27 |
| Low risk  (P_50_ – P_80_] | 1.09 | 1.19 | 2.09 | 6.24 | 1.11 | 1.72 | 2.42 | 6.08 |
| Very low risk  ≤ P_50_ | 0.47 | 0.58 | 0.72 | 2.74 | 0.48 | 0.9 | 0.95 | 2.55 |

##### Table S7 - Analysis of mortality rates per 1000 habitants over the 12 months following MADS assessment itemized by age and sex categories in CHSS.

| MADS  Risk Pyramid Tiers | Mortality  (cases/1k hab.)  Male 0-20 | Mortality  (cases/1k hab.)  Male 20-40 | Mortality  (cases/1k hab.)  Male 40-60 | Mortality  (cases/1k hab.)  Male >60 | Mortality  (cases/1k hab.)  Female 0-20 | Mortality  (cases/1k hab.)  Female 20-40 | Mortality  (cases/1k hab.)  Female 40-60 | Mortality  (cases/1k hab.)  Female >60 |
| --- | --- | --- | --- | --- | --- | --- | --- | --- |
| Very high risk  > P_99_ | 0 | 4.6 | 12.2 | 131 | 0 | 0 | 9.7 | 96.5 |
| High risk  (P_95_ – P_99_] | 0 | 3.2 | 12.3 | 104.8 | 0 | 0 | 4.9 | 70.4 |
| Moderate risk  (P_80_ – P_95_] | 1.1 | 0.6 | 4.7 | 73.3 | 1.2 | 0.2 | 2 | 52.8 |
| Low risk  (P_50_ – P_80_] | 0 | 0.5 | 2.2 | 36.7 | 0 | 0.1 | 1.2 | 29.8 |
| Very low risk  ≤ P_50_ | 0.1 | 0.2 | 1.1 | 14.4 | 0 | 0.1 | 0.8 | 9.3 |

##### Table S8 - Analysis of average pharmacological expenditure per person over the 12 months following MADS assessment itemized by age and sex categories in CHSS.

| MADS  Risk Pyramid Tiers | Medication expenditure in €  (per person)  Male 0-20 | Medication expenditure in €  (per person)  Male 20-40 | Medication expenditure in €  (per person)  Male 40-60 | Medication expenditure in €  (per person)  Male >60 | Medication expenditure in €  (per person)  Female 0-20 | Medication expenditure in €  (per person)  Female 20-40 | Medication expenditure in €  (per person)  Female 40-60 | Medication expenditure in €  (per person)  Female >60 |
| --- | --- | --- | --- | --- | --- | --- | --- | --- |
| Very high risk  > P_99_ | 390 | 1,279 | 1,552 | 1,525 | 464 | 590 | 1,166 | 1,200 |
| High risk  (P_95_ – P_99_] | 350 | 256 | 831 | 1,193 | 160 | 234 | 709 | 919 |
| Moderate risk  (P_80_ – P_95_] | 126 | 148 | 358 | 1,029 | 117 | 153 | 353 | 725 |
| Low risk  (P_50_ – P_80_] | 46 | 85 | 201 | 749 | 34 | 106 | 222 | 510 |
| Very low risk  ≤ P_50_ | 25 | 43 | 77 | 364 | 11 | 41 | 98 | 246 |

##### Table S9 - Analysis of average hospitalization costs per person over the 12 months following MADS assessment itemized by age and sex categories in CHSS.

| MADS  Risk Pyramid Tiers | Hospitalization expenditure in €  (per person)  Male 0-20 | Hospitalization expenditure in €  (per person)  Male 20-40 | Hospitalization expenditure in €  (per person)  Male 40-60 | Hospitalization expenditure in €  (per person)  Male >60 | Hospitalization expenditure in €  (per person)  Female 0-20 | Hospitalization expenditure in €  (per person)  Female 20-40 | Hospitalization expenditure in €  (per person)  Female 40-60 | Hospitalization expenditure in €  (per person)  Female >60 |
| --- | --- | --- | --- | --- | --- | --- | --- | --- |
| Very high risk  > P_99_ | 271 | 466 | 496 | 777 | 489 | 426 | 407 | 645 |
| High risk  (P_95_ – P_99_] | 124 | 156 | 317 | 679 | 298 | 175 | 229 | 496 |
| Moderate risk  (P_80_ – P_95_] | 114 | 93 | 200 | 583 | 122 | 131 | 155 | 388 |
| Low risk  (P_50_ – P_80_] | 66 | 65 | 114 | 385 | 51 | 107 | 103 | 267 |
| Very low risk  ≤ P_50_ | 33 | 32 | 48 | 164 | 23 | 63 | 49 | 107 |

##### Table S10 - Analysis of average total healthcare expenditure per person over the 12 months following MADS assessment itemized by age and sex categories in CHSS.

| MADS  Risk Pyramid Tiers | Total expenditure in €  (per person)  Male 0-20 | Total expenditure in €  (per person)  Male 20-40 | Total expenditure in €  (per person)  Male 40-60 | Total expenditure in €  (per person)  Male >60 | Total expenditure in €  (per person)  Female 0-20 | Total expenditure in €  (per person)  Female 20-40 | Total expenditure in €  (per person)  Female 40-60 | Total expenditure in €  (per person)  Female >60 |
| --- | --- | --- | --- | --- | --- | --- | --- | --- |
| Very high risk  > P_99_ | 6,051 | 10,145 | 12,235 | 17,674 | 10,242 | 7,439 | 11,028 | 14,934 |
| High risk  (P_95_ – P_99_] | 3,532 | 3,431 | 8,477 | 15,225 | 4,669 | 3,256 | 5,776 | 10,047 |
| Moderate risk  (P_80_ – P_95_] | 2,224 | 2,014 | 4,784 | 11,387 | 2,782 | 2,036 | 2,987 | 7,021 |
| Low risk  (P_50_ – P_80_] | 1,350 | 1,308 | 2,677 | 7,392 | 1,191 | 1,517 | 1,822 | 4,513 |
| Very low risk  ≤ P_50_ | 813 | 757 | 1,039 | 3,475 | 721 | 929 | 910 | 1,776 |

##### Table S11- Analysis of average number of different antipsychotics prescribed per person over the 12 months following MADS assessment itemized by age and sex categories in CHSS.

| MADS  Risk Pyramid Tiers | Antipsychotic  (N05A)  (per person)  Male 0-20 | Antipsychotic  (N05A)  (per person)  Male 20-40 | Antipsychotic  (N05A)  (per person)  Male 40-60 | Antipsychotic  (N05A)  (per person)  Male >60 | Antipsychotic  (N05A)  (per person)  Female 0-20 | Antipsychotic  (N05A)  (per person)  Female 20-40 | Antipsychotic  (N05A)  (per person)  Female 40-60 | Antipsychotic  (N05A)  (per person)  Female >60 |
| --- | --- | --- | --- | --- | --- | --- | --- | --- |
| Very high risk  > P_99_ | 1.23 | 1.57 | 1.65 | 1.08 | 0.87 | 0.88 | 0.95 | 0.68 |
| High risk  (P_95_ – P_99_] | 0.74 | 0.27 | 0.73 | 0.20 | 0.33 | 0.18 | 0.28 | 0.17 |
| Moderate risk  (P_80_ – P_95_] | 0.41 | 0.24 | 0.18 | 0.14 | 0.26 | 0.13 | 0.08 | 0.12 |
| Low risk  (P_50_ – P_80_] | 0.17 | 0.15 | 0.16 | 0.09 | 0.09 | 0.07 | 0.08 | 0.09 |
| Very low risk  ≤ P_50_ | 0.10 | 0.09 | 0.09 | 0.06 | 0.06 | 0.06 | 0.06 | 0.05 |

##### Table S12- Analysis of average number of different anxiolytics prescribed per person over the 12 months following MADS assessment itemized by age and sex categories in CHSS.

| MADS  Risk Pyramid Tiers | Anxiolytic  (N05B)  (per person)  Male 0-20 | Anxiolytic  (N05B)  (per person)  Male 20-40 | Anxiolytic  (N05B)  (per person)  Male 40-60 | Anxiolytic  (N05B)  (per person)  Male >60 | Anxiolytic  (N05B)  (per person)  Female 0-20 | Anxiolytic  (N05B)  (per person)  Female 20-40 | Anxiolytic  (N05B)  (per person)  Female 40-60 | Anxiolytic  (N05B)  (per person)  Female >60 |
| --- | --- | --- | --- | --- | --- | --- | --- | --- |
| Very high risk  > P_99_ | 0.24 | 0.37 | 0.74 | 0.45 | 0.18 | 0.41 | 0.72 | 0.67 |
| High risk  (P_95_ – P_99_] | 0.13 | 0.30 | 0.58 | 0.52 | 0.14 | 0.32 | 0.63 | 0.64 |
| Moderate risk  (P_80_ – P_95_] | 0.19 | 0.29 | 0.47 | 0.49 | 0.15 | 0.26 | 0.47 | 0.58 |
| Low risk  (P_50_ – P_80_] | 0.06 | 0.25 | 0.37 | 0.47 | 0.09 | 0.22 | 0.34 | 0.51 |
| Very low risk  ≤ P_50_ | 0.04 | 0.11 | 0.24 | 0.38 | 0.07 | 0.16 | 0.25 | 0.40 |

##### Table S13- Analysis of average number of different hypnotics and sedatives prescribed per person over the 12 months following MADS assessment itemized by age and sex categories in CHSS.

| MADS  Risk Pyramid Tiers | Hypnotics and sedatives  (N05C)  (per person)  Male 0-20 | Hypnotics and sedatives  (N05C)  (per person)  Male 20-40 | Hypnotics and sedatives  (N05C)  (per person)  Male 40-60 | Hypnotics and sedatives  (N05C)  (per person)  Male >60 | Hypnotics and sedatives  (N05C)  (per person)  Female 0-20 | Hypnotics and sedatives  (N05C)  (per person)  Female 20-40 | Hypnotics and sedatives  (N05C)  (per person)  Female 40-60 | Hypnotics and sedatives  (N05C)  (per person)  Female >60 |
| --- | --- | --- | --- | --- | --- | --- | --- | --- |
| Very high risk  > P_99_ | 0.10 | 0.11 | 0.20 | 0.19 | 0.01 | 0.18 | 0.25 | 0.28 |
| High risk  (P_95_ – P_99_] | 0.01 | 0.08 | 0.14 | 0.20 | 0.05 | 0.05 | 0.16 | 0.21 |
| Moderate risk  (P_80_ – P_95_] | 0.02 | 0.05 | 0.10 | 0.16 | 0.04 | 0.04 | 0.10 | 0.18 |
| Low risk  (P_50_ – P_80_] | 0.00 | 0.03 | 0.09 | 0.15 | 0.01 | 0.03 | 0.07 | 0.17 |
| Very low risk  ≤ P_50_ | 0.00 | 0.03 | 0.07 | 0.13 | 0.00 | 0.02 | 0.05 | 0.14 |

##### Table S14- Analysis of average number of different antidepressants prescribed per person over the 12 months following MADS assessment itemized by age and sex categories in CHSS.

| MADS  Risk Pyramid Tiers | Antidepressant  (N06A)  (per person)  Male 0-20 | Antidepressant  (N06A)  (per person)  Male 20-40 | Antidepressant  (N06A)  (per person)  Male 40-60 | Antidepressant  (N06A)  (per person)  Male >60 | Antidepressant  (N06A)  (per person)  Female 0-20 | Antidepressant  (N06A)  (per person)  Female 20-40 | Antidepressant  (N06A)  (per person)  Female 40-60 | Antidepressant  (N06A)  (per person)  Female >60 |
| --- | --- | --- | --- | --- | --- | --- | --- | --- |
| Very high risk  > P_99_ | 0.64 | 0.52 | 0.90 | 0.80 | 0.91 | 0.76 | 1.21 | 1.11 |
| High risk  (P_95_ – P_99_] | 0.46 | 0.56 | 0.78 | 0.77 | 0.64 | 0.63 | 0.91 | 0.94 |
| Moderate risk  (P_80_ – P_95_] | 0.20 | 0.42 | 0.52 | 0.46 | 0.31 | 0.45 | 0.57 | 0.62 |
| Low risk  (P_50_ – P_80_] | 0.05 | 0.27 | 0.32 | 0.22 | 0.10 | 0.27 | 0.34 | 0.34 |
| Very low risk  ≤ P_50_ | 0.05 | 0.15 | 0.16 | 0.16 | 0.08 | 0.18 | 0.17 | 0.22 |

#### 1.2 THL cohort

##### Table S15 - Analysis of mortality rates per 1000 habitants over the 12 months following MADS assessment itemized by age and sex categories in THL.

| MADS  Risk Pyramid Tiers | Mortality  (cases/1k hab.)  Male 0-20 | Mortality  (cases/1k hab.)  Male 20-40 | Mortality  (cases/1k hab.)  Male 40-60 | Mortality  (cases/1k hab.)  Male >60 | Mortality  (cases/1k hab.)  Female 0-20 | Mortality  (cases/1k hab.)  Female 20-40 | Mortality  (cases/1k hab.)  Female 40-60 | Mortality  (cases/1k hab.)  Female >60 |
| --- | --- | --- | --- | --- | --- | --- | --- | --- |
| Very high risk  > P_99_ | N.A. | 0.00 | 50.00 | 55.00 | N.A. | 0.00 | 0.00 | 39.00 |
| High risk  (P_95_ – P_99_] | N.A. | 0.00 | 0.00 | 78.00 | N.A. | 0.00 | 9.00 | 40.00 |
| Moderate risk  (P_80_ – P_95_] | N.A. | 0.00 | 9.00 | 56.00 | N.A. | 0.00 | 5.00 | 35.00 |
| Low risk  (P_50_ – P_80_] | N.A. | 0.00 | 5.00 | 27.00 | N.A. | 0.00 | 3.00 | 17.00 |
| Very low risk  ≤ P_50_ | N.A. | 2.00 | 3.00 | 14.00 | N.A. | 0.00 | 1.00 | 9.00 |

##### Table S16 - Analysis of average pharmacological expenditure per person over the 12 months following MADS assessment itemized by age and sex categories in THL.

| MADS  Risk Pyramid Tiers | Medication expenditure in €  (per person)  Male 0-20 | Medication expenditure in €  (per person)  Male 20-40 | Medication expenditure in €  (per person)  Male 40-60 | Medication expenditure in €  (per person)  Male >60 | Medication expenditure in €  (per person)  Female 0-20 | Medication expenditure in €  (per person)  Female 20-40 | Medication expenditure in €  (per person)  Female 40-60 | Medication expenditure in €  (per person)  Female >60 |
| --- | --- | --- | --- | --- | --- | --- | --- | --- |
| Very high risk  > P_99_ | N.A. | 277 | 1,012 | 1,013 | N.A. | 501 | 601 | 1,125 |
| High risk  (P_95_ – P_99_] | N.A. | 1,550 | 864 | 1,177 | N.A. | 482 | 1,074 | 1,344 |
| Moderate risk  (P_80_ – P_95_] | N.A. | 863 | 925 | 1,349 | N.A. | 313 | 733 | 1,123 |
| Low risk  (P_50_ – P_80_] | N.A. | 169 | 588 | 1,079 | N.A. | 219 | 608 | 879 |
| Very low risk  ≤ P_50_ | N.A. | 84 | 182 | 564 | N.A. | 148 | 207 | 435 |

##### Table S18 - Analysis of average hospitalization costs per person over the 12 months following MADS assessment itemized by age and sex categories in THL.

| MADS  Risk Pyramid Tiers | Hospitalization expenditure in €  (per person)  Male 0-20 | Hospitalization expenditure in €  (per person)  Male 20-40 | Hospitalization expenditure in €  (per person)  Male 40-60 | Hospitalization expenditure in €  (per person)  Male >60 | Hospitalization expenditure in €  (per person)  Female 0-20 | Hospitalization expenditure in €  (per person)  Female 20-40 | Hospitalization expenditure in €  (per person)  Female 40-60 | Hospitalization expenditure in €  (per person)  Female >60 |
| --- | --- | --- | --- | --- | --- | --- | --- | --- |
| Very high risk  > P_99_ | N.A. | 198 | 139 | 412 | N.A. | 305 | 272 | 221 |
| High risk  (P_95_ – P_99_] | N.A. | 837 | 247 | 308 | N.A. | 716 | 417 | 255 |
| Moderate risk  (P_80_ – P_95_] | N.A. | 245 | 90 | 369 | N.A. | 160 | 193 | 237 |
| Low risk  (P_50_ – P_80_] | N.A. | 19 | 94 | 207 | N.A. | 174 | 160 | 227 |
| Very low risk  ≤ P_50_ | N.A. | 28 | 115 | 160 | N.A. | 71 | 105 | 121 |

##### Table S19- Analysis of average number of different antipsychotics prescribed per person over the 12 months following MADS assessment itemized by age and sex categories in THL.

| MADS  Risk Pyramid Tiers | Antipsychotic  (N05A)  (per person)  Male 0-20 | Antipsychotic  (N05A)  (per person)  Male 20-40 | Antipsychotic  (N05A)  (per person)  Male 40-60 | Antipsychotic  (N05A)  (per person)  Male >60 | Antipsychotic  (N05A)  (per person)  Female 0-20 | Antipsychotic  (N05A)  (per person)  Female 20-40 | Antipsychotic  (N05A)  (per person)  Female 40-60 | Antipsychotic  (N05A)  (per person)  Female >60 |
| --- | --- | --- | --- | --- | --- | --- | --- | --- |
| Very high risk  > P_99_ | N.A. | 0.63 | 0.70 | 0.58 | N.A. | 0.38 | 0.52 | 0.62 |
| High risk  (P_95_ – P_99_] | N.A. | 0.06 | 0.28 | 0.27 | N.A. | 0.11 | 0.30 | 0.28 |
| Moderate risk  (P_80_ – P_95_] | N.A. | 0.06 | 0.07 | 0.09 | N.A. | 0.06 | 0.07 | 0.09 |
| Low risk  (P_50_ – P_80_] | N.A. | 0.00 | 0.03 | 0.03 | N.A. | 0.02 | 0.02 | 0.04 |
| Very low risk  ≤ P_50_ | N.A. | 0.00 | 0.01 | 0.02 | N.A. | 0.01 | 0.01 | 0.02 |

##### Table S20- Analysis of average number of different anxiolytics prescribed per person over the 12 months following MADS assessment itemized by age and sex categories in THL.

| MADS  Risk Pyramid Tiers | Anxiolytic  (N05B)  (per person)  Male 0-20 | Anxiolytic  (N05B)  (per person)  Male 20-40 | Anxiolytic  (N05B)  (per person)  Male 40-60 | Anxiolytic  (N05B)  (per person)  Male >60 | Anxiolytic  (N05B)  (per person)  Female 0-20 | Anxiolytic  (N05B)  (per person)  Female 20-40 | Anxiolytic  (N05B)  (per person)  Female 40-60 | Anxiolytic  (N05B)  (per person)  Female >60 |
| --- | --- | --- | --- | --- | --- | --- | --- | --- |
| Very high risk  > P_99_ | N.A. | 0.13 | 0.30 | 0.20 | N.A. | 0.13 | 0.24 | 0.20 |
| High risk  (P_95_ – P_99_] | N.A. | 0.19 | 0.16 | 0.16 | N.A. | 0.22 | 0.21 | 0.20 |
| Moderate risk  (P_80_ – P_95_] | N.A. | 0.04 | 0.06 | 0.08 | N.A. | 0.07 | 0.08 | 0.10 |
| Low risk  (P_50_ – P_80_] | N.A. | 0.02 | 0.02 | 0.05 | N.A. | 0.04 | 0.03 | 0.06 |
| Very low risk  ≤ P_50_ | N.A. | 0.01 | 0.01 | 0.02 | N.A. | 0.02 | 0.02 | 0.03 |

##### Table S21- Analysis of average number of different hypnotics and sedatives prescribed per person over the 12 months following MADS assessment itemized by age and sex categories in THL.

| MADS  Risk Pyramid Tiers | Hypnotics and sedatives  (N05C)  (per person)  Male 0-20 | Hypnotics and sedatives  (N05C)  (per person)  Male 20-40 | Hypnotics and sedatives  (N05C)  (per person)  Male 40-60 | Hypnotics and sedatives  (N05C)  (per person)  Male >60 | Hypnotics and sedatives  (N05C)  (per person)  Female 0-20 | Hypnotics and sedatives  (N05C)  (per person)  Female 20-40 | Hypnotics and sedatives  (N05C)  (per person)  Female 40-60 | Hypnotics and sedatives  (N05C)  (per person)  Female >60 |
| --- | --- | --- | --- | --- | --- | --- | --- | --- |
| Very high risk  > P_99_ | N.A. | 0.00 | 0.10 | 0.09 | N.A. | 0.25 | 0.10 | 0.19 |
| High risk  (P_95_ – P_99_] | N.A. | 0.06 | 0.10 | 0.10 | N.A. | 0.07 | 0.14 | 0.15 |
| Moderate risk  (P_80_ – P_95_] | N.A. | 0.03 | 0.04 | 0.10 | N.A. | 0.03 | 0.10 | 0.13 |
| Low risk  (P_50_ – P_80_] | N.A. | 0.01 | 0.04 | 0.06 | N.A. | 0.03 | 0.04 | 0.10 |
| Very low risk  ≤ P_50_ | N.A. | 0.01 | 0.03 | 0.04 | N.A. | 0.00 | 0.03 | 0.07 |

##### Table S22- Analysis of average number of different antidepressants prescribed per person over the 12 months following MADS assessment itemized by age and sex categories in THL.

| MADS  Risk Pyramid Tiers | Antidepressant  (N06A)  (per person)  Male 0-20 | Antidepressant  (N06A)  (per person)  Male 20-40 | Antidepressant  (N06A)  (per person)  Male 40-60 | Antidepressant  (N06A)  (per person)  Male >60 | Antidepressant  (N06A)  (per person)  Female 0-20 | Antidepressant  (N06A)  (per person)  Female 20-40 | Antidepressant  (N06A)  (per person)  Female 40-60 | Antidepressant  (N06A)  (per person)  Female >60 |
| --- | --- | --- | --- | --- | --- | --- | --- | --- |
| Very high risk  > P_99_ | N.A. | 0.38 | 0.50 | 0.42 | N.A. | 0.75 | 0.52 | 0.37 |
| High risk  (P_95_ – P_99_] | N.A. | 0.44 | 0.44 | 0.33 | N.A. | 0.39 | 0.49 | 0.41 |
| Moderate risk  (P_80_ – P_95_] | N.A. | 0.22 | 0.23 | 0.22 | N.A. | 0.22 | 0.38 | 0.29 |
| Low risk  (P_50_ – P_80_] | N.A. | 0.07 | 0.08 | 0.09 | N.A. | 0.11 | 0.13 | 0.13 |
| Very low risk  ≤ P_50_ | N.A. | 0.04 | 0.04 | 0.04 | N.A. | 0.06 | 0.07 | 0.08 |

#### 1.3 UKB cohort

##### Table S23 - Analysis of average number of different pharmacological prescriptions per person over the 12 months following MADS assessment itemized by age and sex categories in UKB.

| MADS  Risk Pyramid Tiers | Number of prescriptions  (per person)  Male 0-20 | Number of prescriptions  (per person)  Male 20-40 | Number of prescriptions  (per person)  Male 40-60 | Number of prescriptions  (per person)  Male >60 | Number of prescriptions  (per person)  Female 0-20 | Number of prescriptions  (per person)  Female 20-40 | Number of prescriptions  (per person)  Female 40-60 | Number of prescriptions  (per person)  Female >60 |
| --- | --- | --- | --- | --- | --- | --- | --- | --- |
| Very high risk  > P_99_ | N.A. | N.A. | 13.63 | 13.43 | N.A. | N.A. | 13.63 | 13.43 |
| High risk  (P_95_ – P_99_] | N.A. | N.A. | 11.91 | 12.20 | N.A. | N.A. | 11.91 | 12.20 |
| Moderate risk  (P_80_ – P_95_] | N.A. | N.A. | 10.76 | 12.29 | N.A. | N.A. | 10.76 | 12.29 |
| Low risk  (P_50_ – P_80_] | N.A. | N.A. | 9.18 | 11.65 | N.A. | N.A. | 9.18 | 11.65 |
| Very low risk  ≤ P_50_ | N.A. | N.A. | 7.11 | 9.88 | N.A. | N.A. | 7.11 | 9.88 |

##### Table S24- Analysis of average number of different antipsychotics prescribed per person over the 12 months following MADS assessment itemized by age and sex categories in UKB.

| MADS  Risk Pyramid Tiers | Antipsychotic  (N05A)  (per person)  Male 0-20 | Antipsychotic  (N05A)  (per person)  Male 20-40 | Antipsychotic  (N05A)  (per person)  Male 40-60 | Antipsychotic  (N05A)  (per person)  Male >60 | Antipsychotic  (N05A)  (per person)  Female 0-20 | Antipsychotic  (N05A)  (per person)  Female 20-40 | Antipsychotic  (N05A)  (per person)  Female 40-60 | Antipsychotic  (N05A)  (per person)  Female >60 |
| --- | --- | --- | --- | --- | --- | --- | --- | --- |
| Very high risk  > P_99_ | N.A. | N.A. | 0.43 | 0.23 | N.A. | N.A. | 0.37 | 0.32 |
| High risk  (P_95_ – P_99_] | N.A. | N.A. | 0.13 | 0.17 | N.A. | N.A. | 0.21 | 0.17 |
| Moderate risk  (P_80_ – P_95_] | N.A. | N.A. | 0.10 | 0.14 | N.A. | N.A. | 0.17 | 0.17 |
| Low risk  (P_50_ – P_80_] | N.A. | N.A. | 0.07 | 0.12 | N.A. | N.A. | 0.12 | 0.17 |
| Very low risk  ≤ P_50_ | N.A. | N.A. | 0.05 | 0.11 | N.A. | N.A. | 0.12 | 0.14 |

##### Table S25- Analysis of average number of different anxiolytics prescribed per person over the 12 months following MADS assessment itemized by age and sex categories in UKB.

| MADS  Risk Pyramid Tiers | Anxiolytic  (N05B)  (per person)  Male 0-20 | Anxiolytic  (N05B)  (per person)  Male 20-40 | Anxiolytic  (N05B)  (per person)  Male 40-60 | Anxiolytic  (N05B)  (per person)  Male >60 | Anxiolytic  (N05B)  (per person)  Female 0-20 | Anxiolytic  (N05B)  (per person)  Female 20-40 | Anxiolytic  (N05B)  (per person)  Female 40-60 | Anxiolytic  (N05B)  (per person)  Female >60 |
| --- | --- | --- | --- | --- | --- | --- | --- | --- |
| Very high risk  > P_99_ | N.A. | N.A. | 0.23 | 0.24 | N.A. | N.A. | 0.33 | 0.26 |
| High risk  (P_95_ – P_99_] | N.A. | N.A. | 0.19 | 0.20 | N.A. | N.A. | 0.20 | 0.19 |
| Moderate risk  (P_80_ – P_95_] | N.A. | N.A. | 0.16 | 0.16 | N.A. | N.A. | 0.17 | 0.15 |
| Low risk  (P_50_ – P_80_] | N.A. | N.A. | 0.09 | 0.13 | N.A. | N.A. | 0.13 | 0.13 |
| Very low risk  ≤ P_50_ | N.A. | N.A. | 0.10 | 0.08 | N.A. | N.A. | 0.09 | 0.10 |

##### Table S26- Analysis of average number of different hypnotics and sedatives prescribed per person over the 12 months following MADS assessment itemized by age and sex categories in UKB.

| MADS  Risk Pyramid Tiers | Hypnotics and sedatives  (N05C)  (per person)  Male 0-20 | Hypnotics and sedatives  (N05C)  (per person)  Male 20-40 | Hypnotics and sedatives  (N05C)  (per person)  Male 40-60 | Hypnotics and sedatives  (N05C)  (per person)  Male >60 | Hypnotics and sedatives  (N05C)  (per person)  Female 0-20 | Hypnotics and sedatives  (N05C)  (per person)  Female 20-40 | Hypnotics and sedatives  (N05C)  (per person)  Female 40-60 | Hypnotics and sedatives  (N05C)  (per person)  Female >60 |
| --- | --- | --- | --- | --- | --- | --- | --- | --- |
| Very high risk  > P_99_ | N.A. | N.A. | 0.18 | 0.27 | N.A. | N.A. | 0.21 | 0.29 |
| High risk  (P_95_ – P_99_] | N.A. | N.A. | 0.16 | 0.21 | N.A. | N.A. | 0.17 | 0.20 |
| Moderate risk  (P_80_ – P_95_] | N.A. | N.A. | 0.17 | 0.21 | N.A. | N.A. | 0.16 | 0.16 |
| Low risk  (P_50_ – P_80_] | N.A. | N.A. | 0.10 | 0.18 | N.A. | N.A. | 0.11 | 0.12 |
| Very low risk  ≤ P_50_ | N.A. | N.A. | 0.08 | 0.10 | N.A. | N.A. | 0.09 | 0.11 |

##### Table S27- Analysis of average number of different antidepressants prescribed per person over the 12 months following MADS assessment itemized by age and sex categories in UKB.

| MADS  Risk Pyramid Tiers | Antidepressant  (N06A)  (per person)  Male 0-20 | Antidepressant  (N06A)  (per person)  Male 20-40 | Antidepressant  (N06A)  (per person)  Male 40-60 | Antidepressant  (N06A)  (per person)  Male >60 | Antidepressant  (N06A)  (per person)  Female 0-20 | Antidepressant  (N06A)  (per person)  Female 20-40 | Antidepressant  (N06A)  (per person)  Female 40-60 | Antidepressant  (N06A)  (per person)  Female >60 |
| --- | --- | --- | --- | --- | --- | --- | --- | --- |
| Very high risk  > P_99_ | N.A. | N.A. | 1.07 | 0.61 | N.A. | N.A. | 0.82 | 0.77 |
| High risk  (P_95_ – P_99_] | N.A. | N.A. | 0.72 | 0.64 | N.A. | N.A. | 0.86 | 0.63 |
| Moderate risk  (P_80_ – P_95_] | N.A. | N.A. | 0.59 | 0.42 | N.A. | N.A. | 0.68 | 0.51 |
| Low risk  (P_50_ – P_80_] | N.A. | N.A. | 0.31 | 0.29 | N.A. | N.A. | 0.49 | 0.37 |
| Very low risk  ≤ P_50_ | N.A. | N.A. | 0.22 | 0.22 | N.A. | N.A. | 0.33 | 0.28 |

### Annex 2: Longitudinal analysis of disease prevalence and incidence of new disease onsets

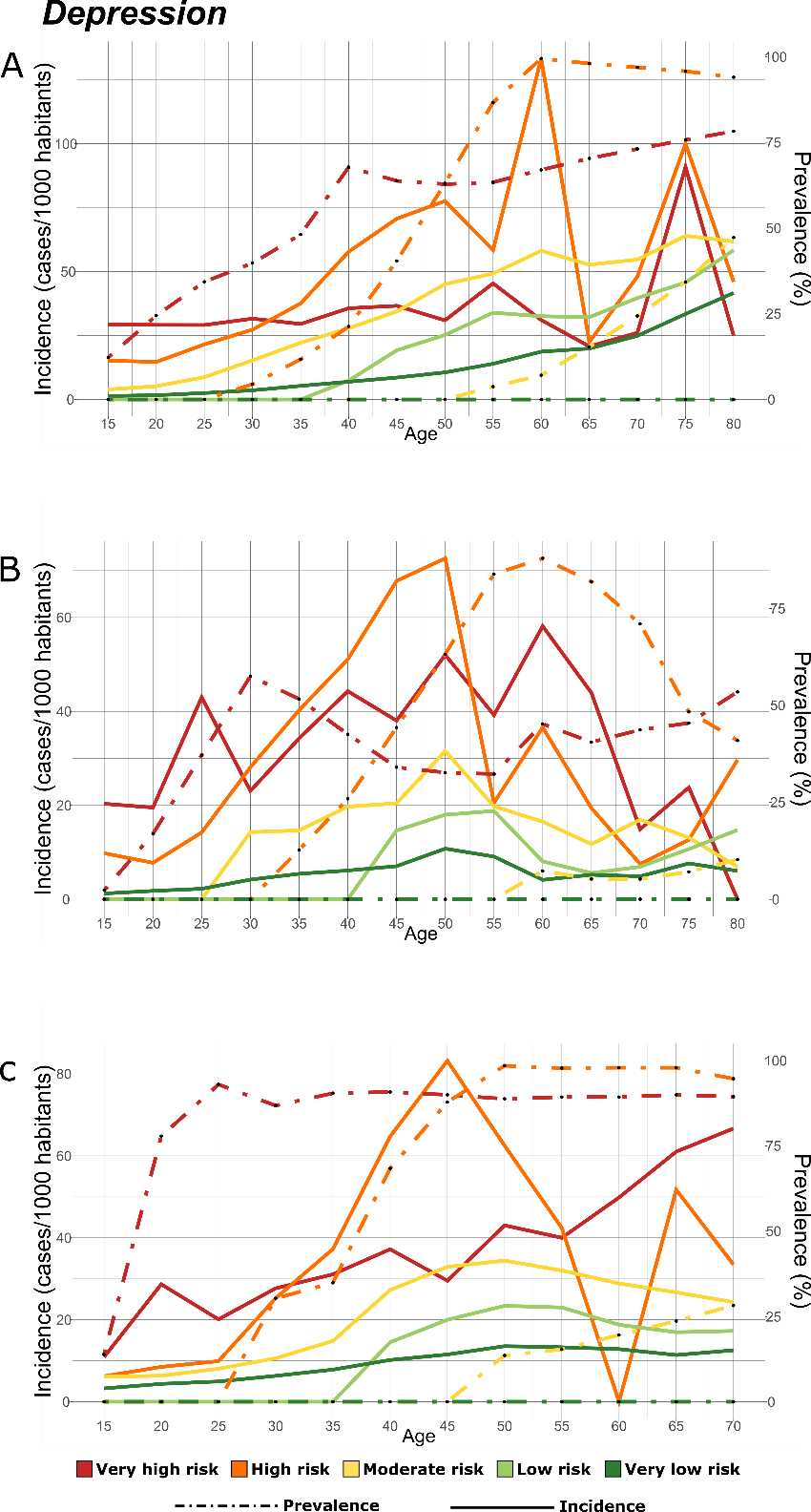

Figure S1 - Longitudinal analysis of disease prevalence and incidence of new onsets of Major Depressive Disorder (ICD-10-CM: F32). *Panel A) CHSS cohort; Panel B) THL cohort; Panel C)UKB cohort. Disease incidence is assessed in a 5-years interval, plotted in the left y-axis and represented with solid lines. Disease prevalence is plotted in the right y-axis and represented with dashed lines. The line colours correspond to the MADS risk pyramid tiers: red: very high-risk group; orange: high-risk group; yellow: medium-risk group; light green: low-risk group; green: very low-risk group.*

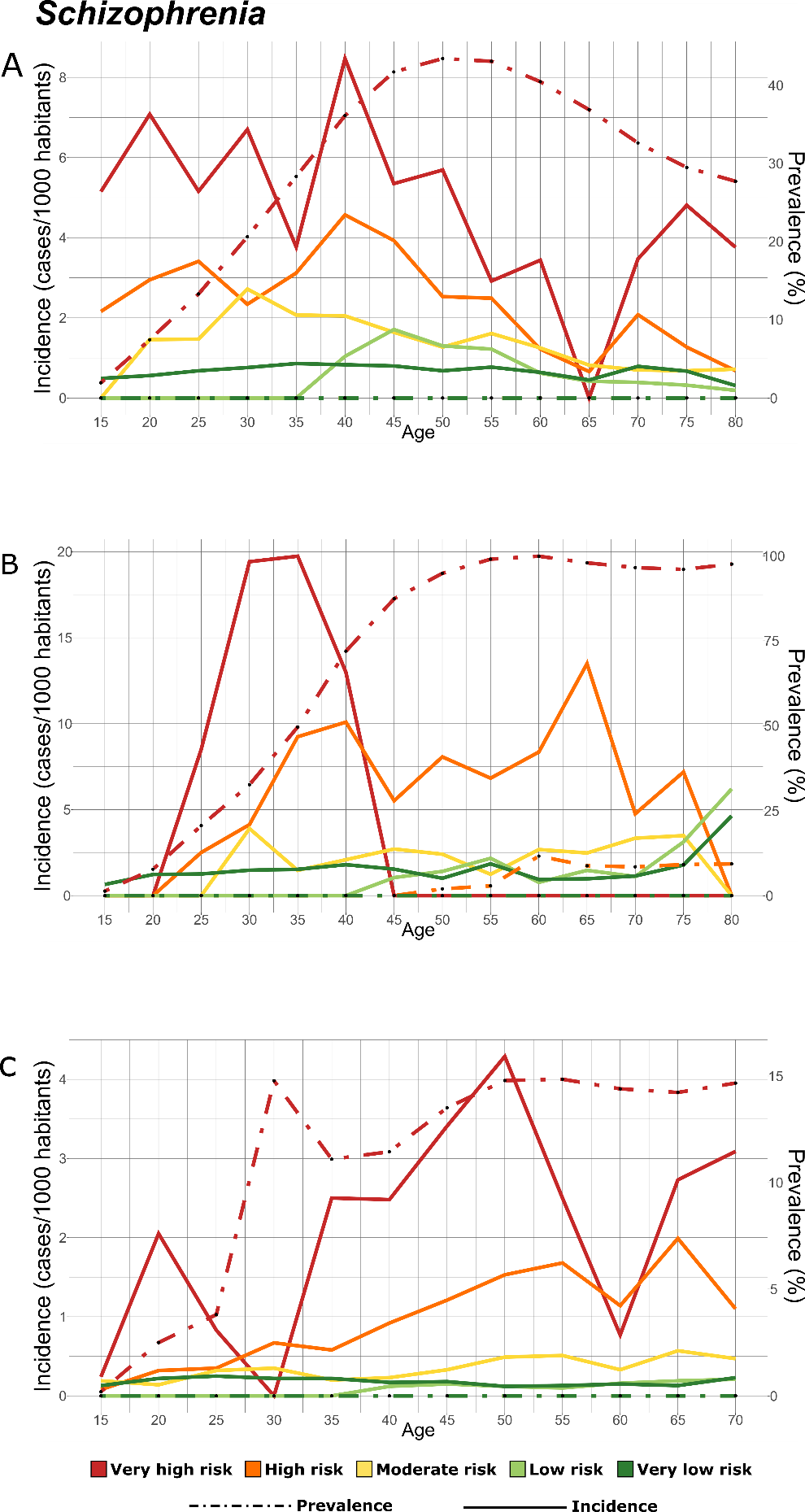

Figure S2 - Longitudinal analysis of disease prevalence and incidence of new onsets of schizophrenia (ICD-10-CM: F20). *Panel A) CHSS cohort; Panel B) THL cohort; Panel C)UKB cohort. Disease incidence is assessed in a 5-years interval, plotted in the left y-axis and represented with solid lines. Disease prevalence is plotted in the right y-axis and represented with dashed lines. The line colours correspond to the MADS risk pyramid tiers: red: very high-risk group; orange: high-risk group; yellow: medium-risk group; light green: low-risk group; green: very low-risk group.*

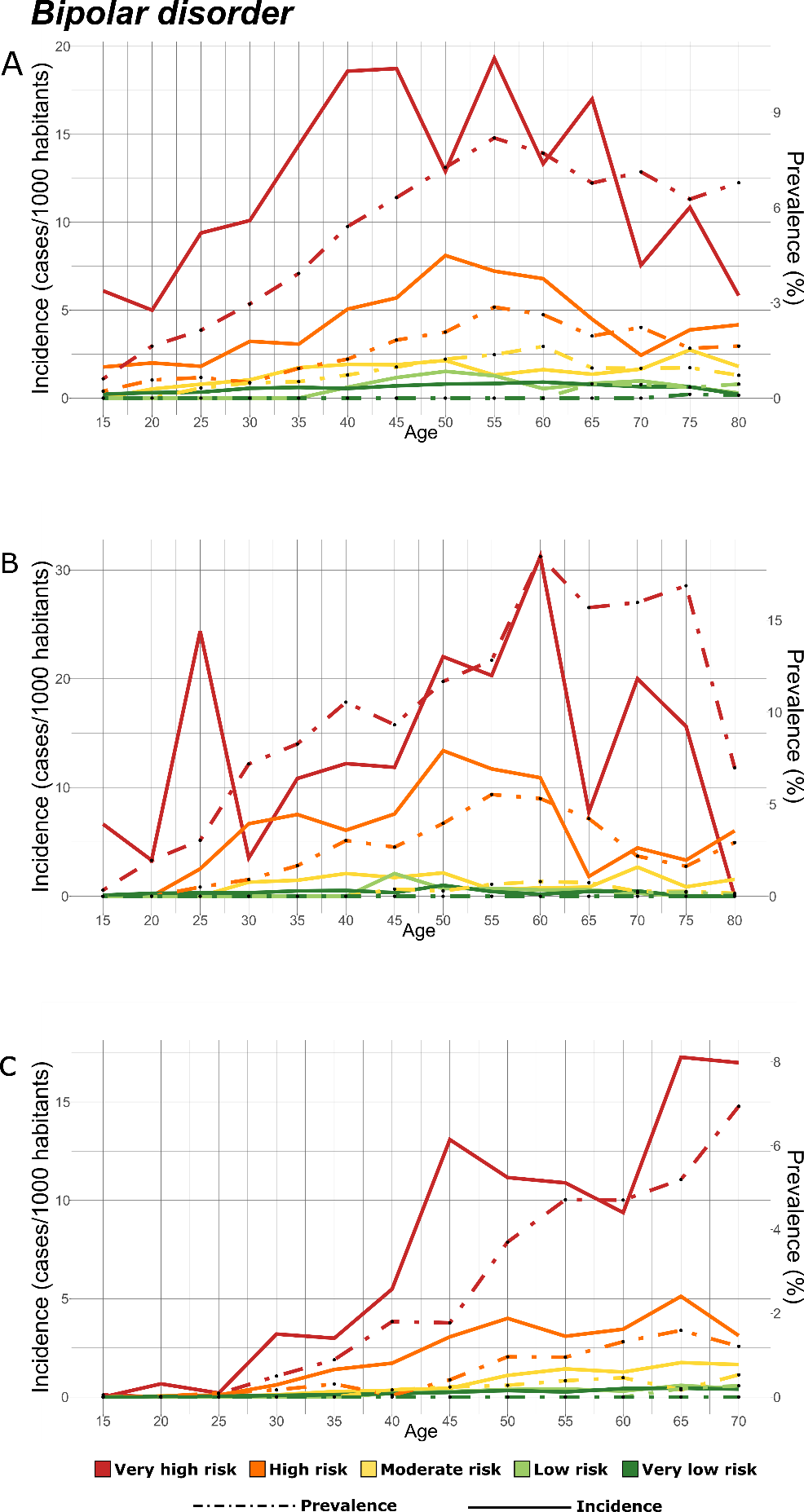

Figure S3 - Longitudinal analysis of disease prevalence and incidence of new onsets of bipolar disorder (ICD-10-CM: F31). *Panel A) CHSS cohort; Panel B) THL cohort; Panel C)UKB cohort. Disease incidence is assessed in a 5-years interval, plotted in the left y-axis and represented with solid lines. Disease prevalence is plotted in the right y-axis and represented with dashed lines. The line colours correspond to the MADS risk pyramid tiers: red: very high-risk group; orange: high-risk group; yellow: medium-risk group; light green: low-risk group; green: very low-risk group.*

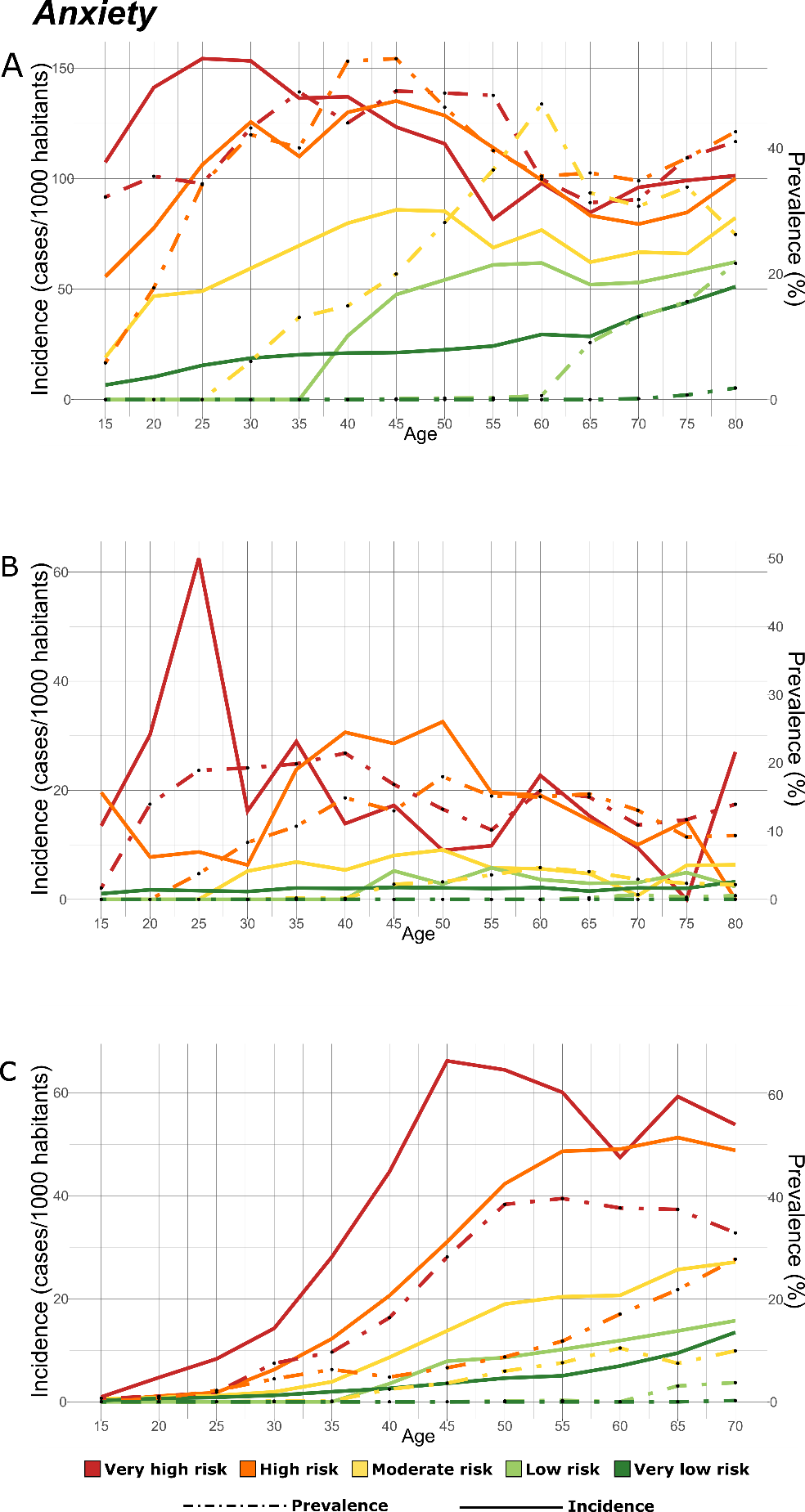

Figure S4 - Longitudinal analysis of disease prevalence and incidence of new onsets of anxiety related disorders (ICD-10-CM: F40-41). *Panel A) CHSS cohort; Panel B) THL cohort; Panel C)UKB cohort. Disease incidence is assessed in a 5-years interval, plotted in the left y-axis and represented with solid lines. Disease prevalence is plotted in the right y-axis and represented with dashed lines. The line colours correspond to the MADS risk pyramid tiers: red: very high-risk group; orange: high-risk group; yellow: medium-risk group; light green: low-risk group; green: very low-risk group.*

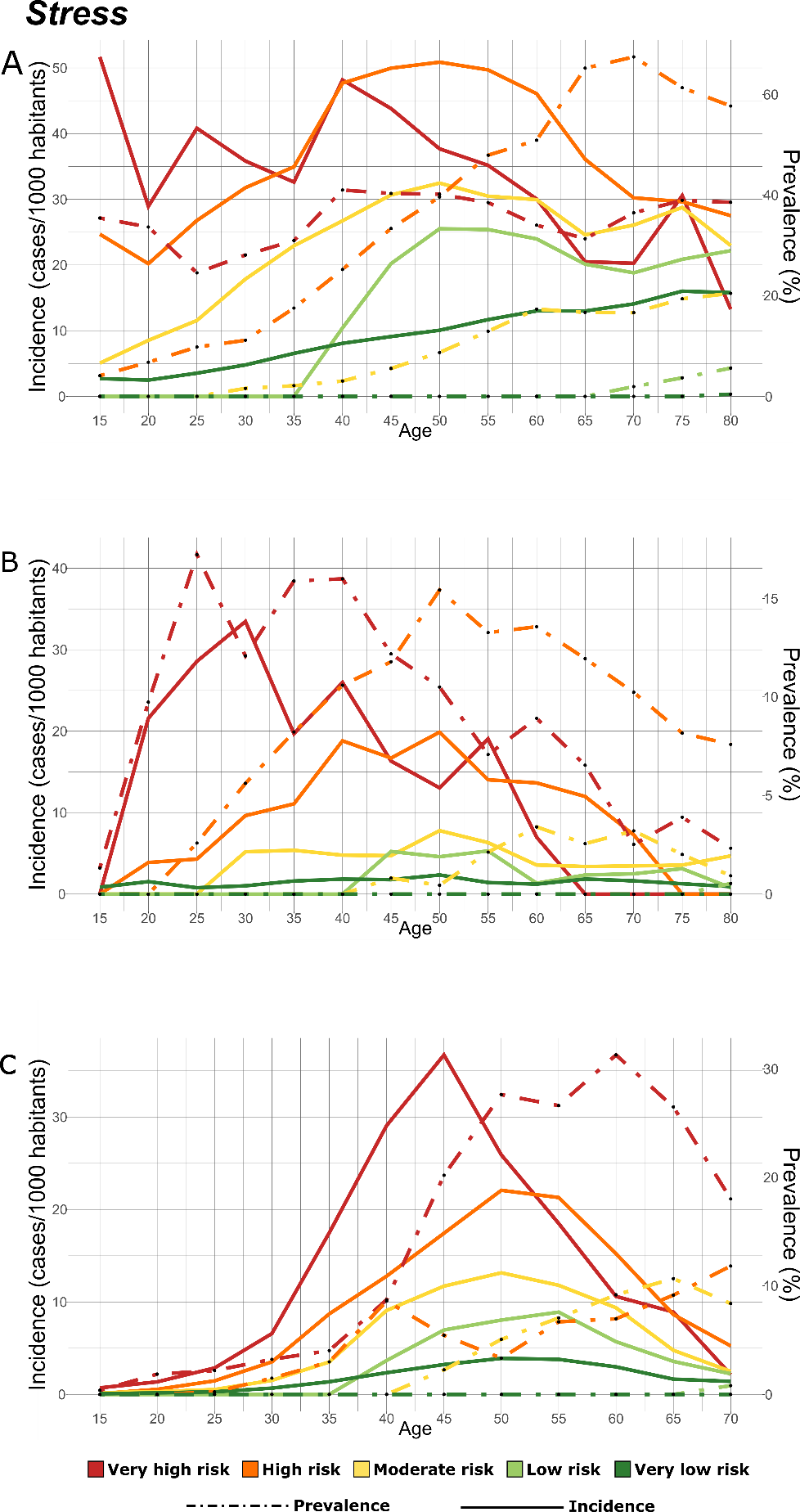

Figure S5 - Longitudinal analysis of disease prevalence and incidence of new onsets of stress related disorders (ICD-10-CM: F43). *Panel A) CHSS cohort; Panel B) THL cohort; Panel C)UKB cohort. Disease incidence is assessed in a 5-years interval, plotted in the left y-axis and represented with solid lines. Disease prevalence is plotted in the right y-axis and represented with dashed lines. The line colours correspond to the MADS risk pyramid tiers: red: very high-risk group; orange: high-risk group; yellow: medium-risk group; light green: low-risk group; green: very low-risk group.*

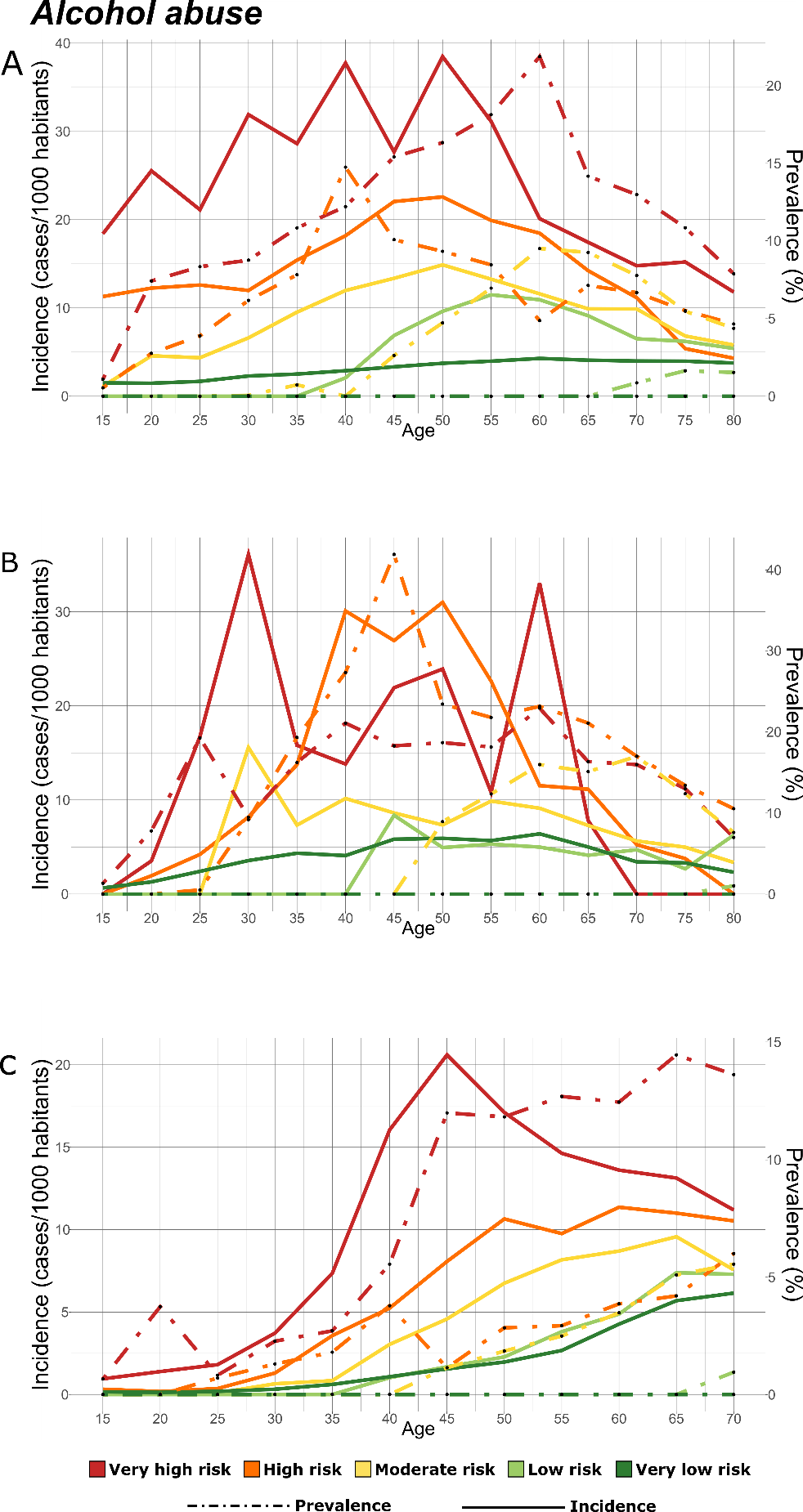

Figure S6 - Longitudinal analysis of disease prevalence and incidence of new onsets of mental disorders related to alcohol abuse (ICD-10-CM: F10). *Panel A) CHSS cohort; Panel B) THL cohort; Panel C)UKB cohort. Disease incidence is assessed in a 5-years interval, plotted in the left y-axis and represented with solid lines. Disease prevalence is plotted in the right y-axis and represented with dashed lines. The line colours correspond to the MADS risk pyramid tiers: red: very high-risk group; orange: high-risk group; yellow: medium-risk group; light green: low-risk group; green: very low-risk group.*

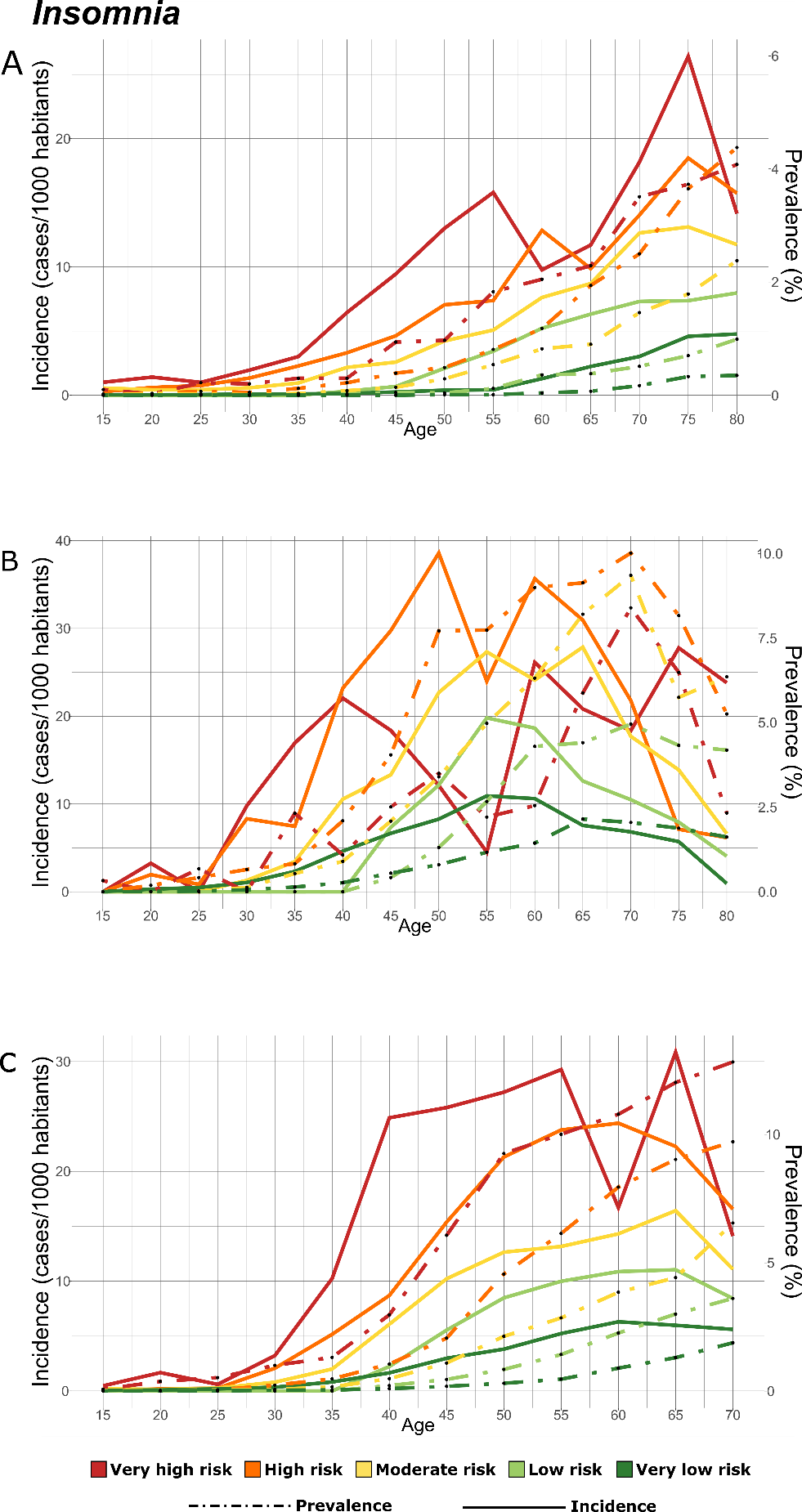

Figure S7 - Longitudinal analysis of disease prevalence and incidence of new onsets of insomnia (ICD-10-CM: G47). *Panel A) CHSS cohort; Panel B) THL cohort; Panel C)UKB cohort. Disease incidence is assessed in a 5-years interval, plotted in the left y-axis and represented with solid lines. Disease prevalence is plotted in the right y-axis and represented with dashed lines. The line colours correspond to the MADS risk pyramid tiers: red: very high-risk group; orange: high-risk group; yellow: medium-risk group; light green: low-risk group; green: very low-risk group.*

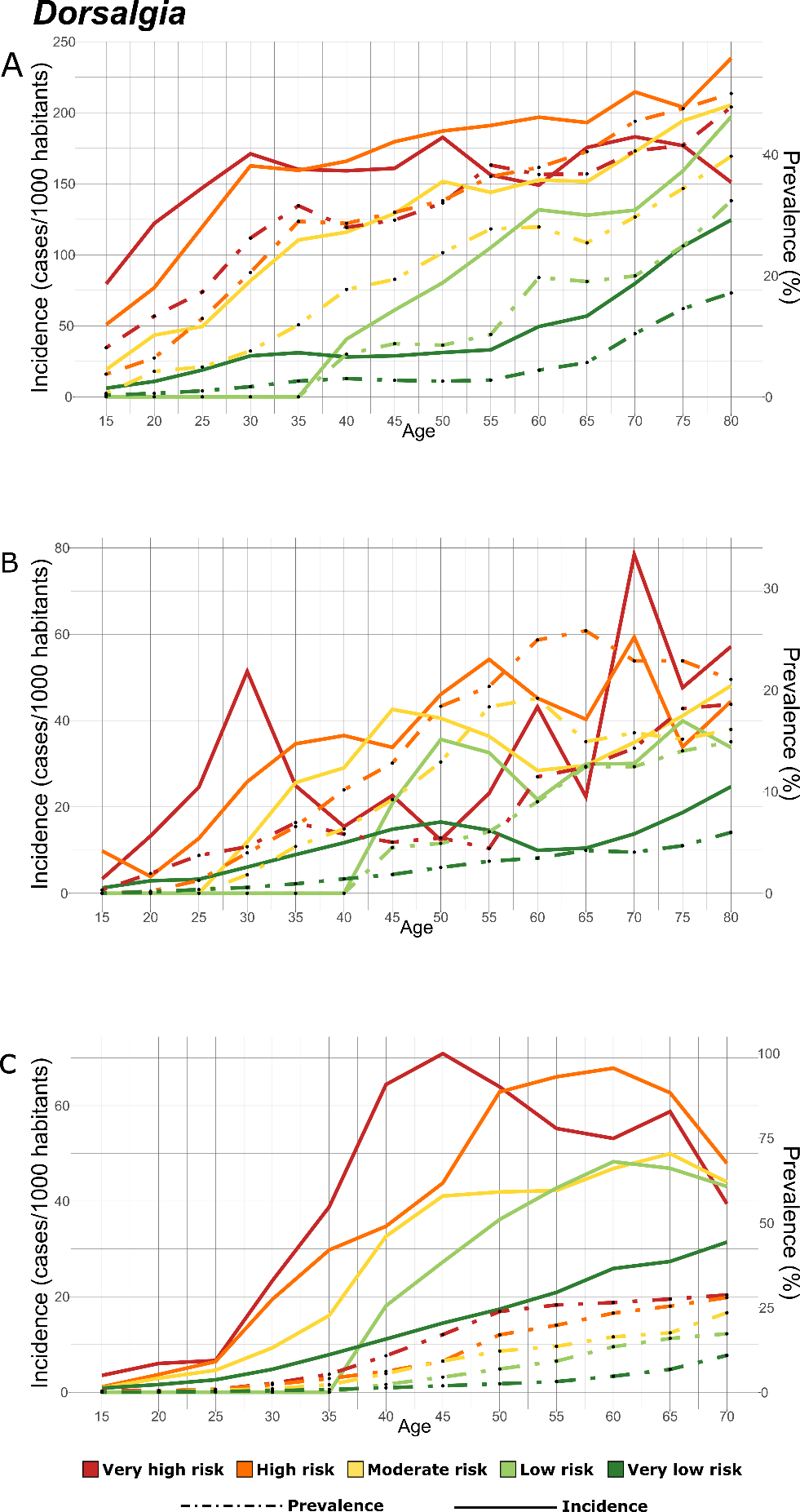

Figure S8 - Longitudinal analysis of disease prevalence and incidence of new onsets of dorsalgia (ICD-10-CM: M54). *Panel A) CHSS cohort; Panel B) THL cohort; Panel C)UKB cohort. Disease incidence is assessed in a 5-years interval, plotted in the left y-axis and represented with solid lines. Disease prevalence is plotted in the right y-axis and represented with dashed lines. The line colours correspond to the MADS risk pyramid tiers: red: very high-risk group; orange: high-risk group; yellow: medium-risk group; light green: low-risk group; green: very low-risk group.*

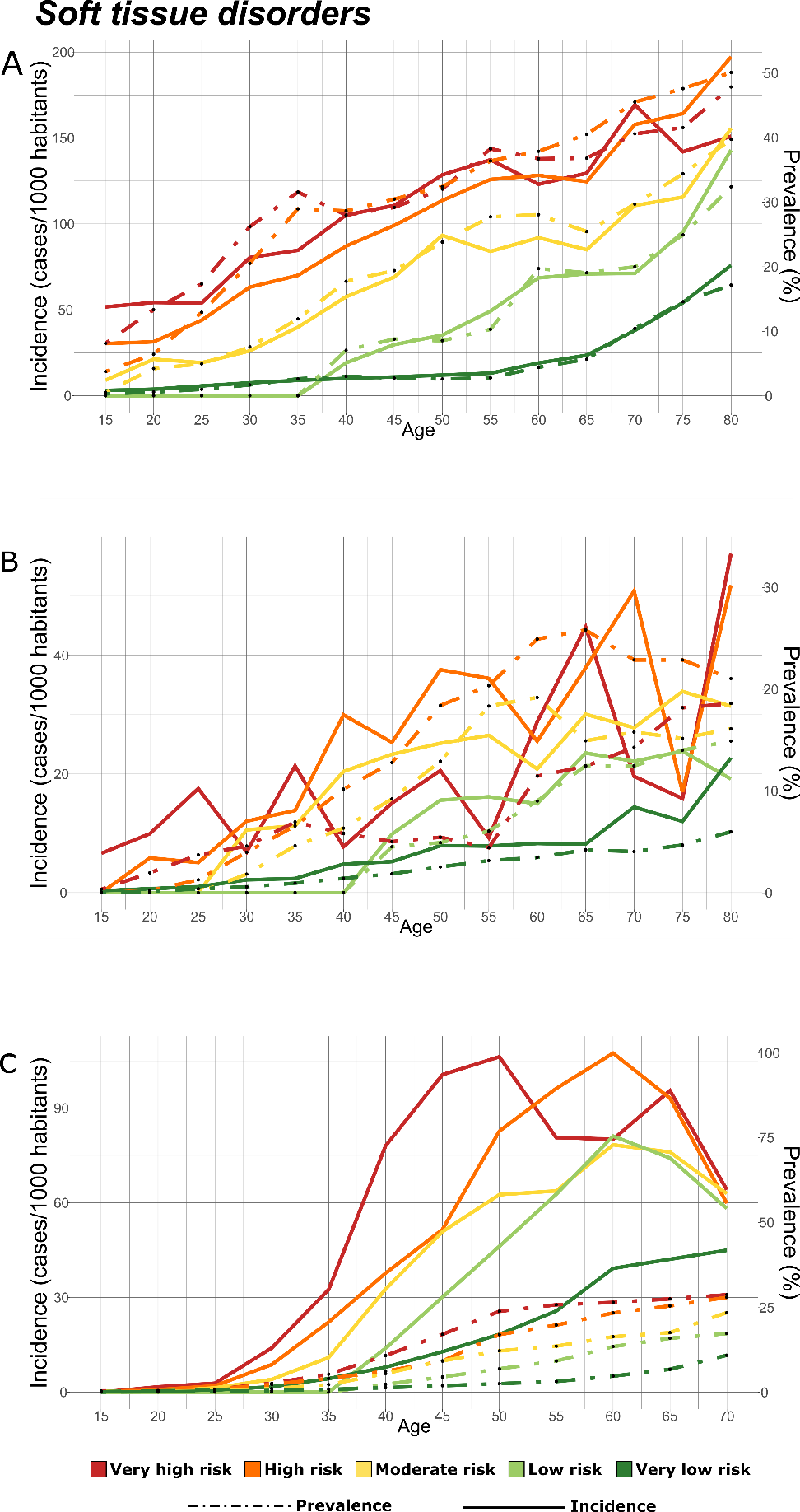

Figure S9 - Longitudinal analysis of disease prevalence and incidence of new onsets of soft tissue disorders not classified elsewhere (ICD-10-CM: M79). *Panel A) CHSS cohort; Panel B) THL cohort; Panel C)UKB cohort. Disease incidence is assessed in a 5-years interval, plotted in the left y-axis and represented with solid lines. Disease prevalence is plotted in the right y-axis and represented with dashed lines. The line colours correspond to the MADS risk pyramid tiers: red: very high-risk group; orange: high-risk group; yellow: medium-risk group; light green: low-risk group; green: very low-risk group.*

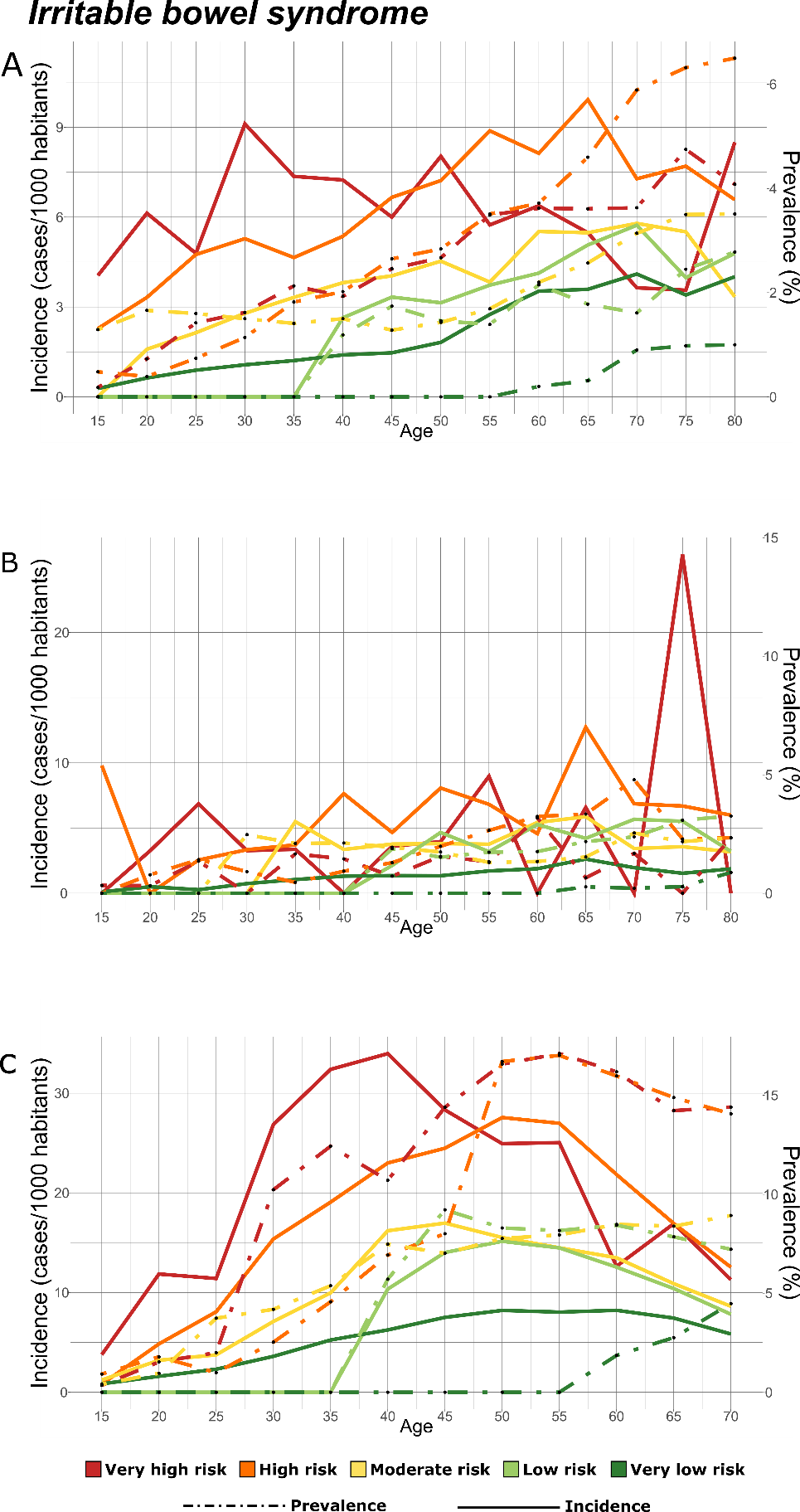

Figure S10 - Longitudinal analysis of disease prevalence and incidence of new onsets of irritable bowel syndrome (ICD-10-CM: K58). *Panel A) CHSS cohort; Panel B) THL cohort; Panel C)UKB cohort. Disease incidence is assessed in a 5-years interval, plotted in the left y-axis and represented with solid lines. Disease prevalence is plotted in the right y-axis and represented with dashed lines. The line colours correspond to the MADS risk pyramid tiers: red: very high-risk group; orange: high-risk group; yellow: medium-risk group; light green: low-risk group; green: very low-risk group.*

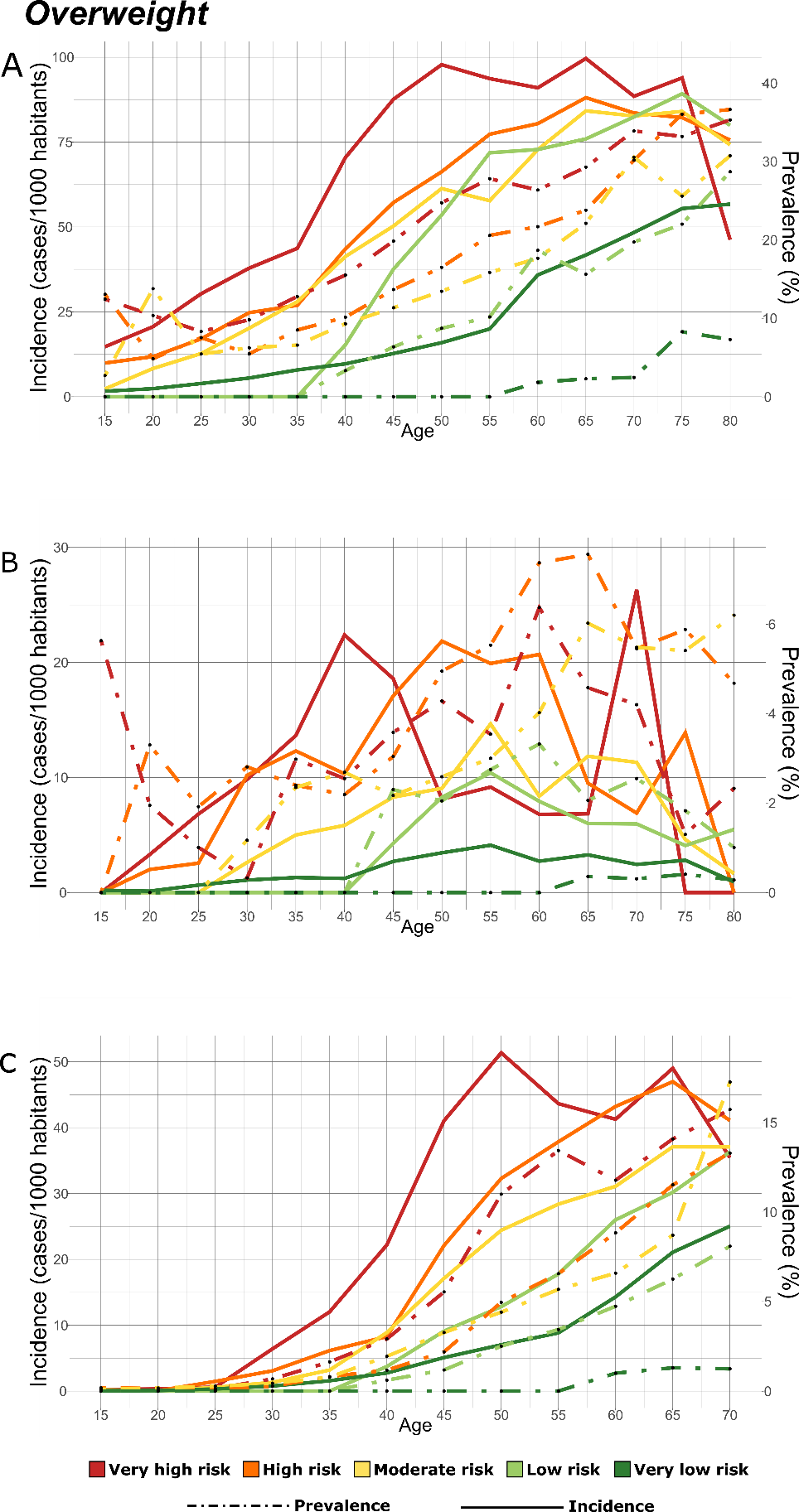

Figure S11 - Longitudinal analysis of disease prevalence and incidence of new onsets of overweight and obesity MDD (ICD-10-CM: E66). *Panel A) CHSS cohort; Panel B) THL cohort; Panel C)UKB cohort. Disease incidence is assessed in a 5-years interval, plotted in the left y-axis and represented with solid lines. Disease prevalence is plotted in the right y-axis and represented with dashed lines. The line colours correspond to the MADS risk pyramid tiers: red: very high-risk group; orange: high-risk group; yellow: medium-risk group; light green: low-risk group; green: very low-risk group.*

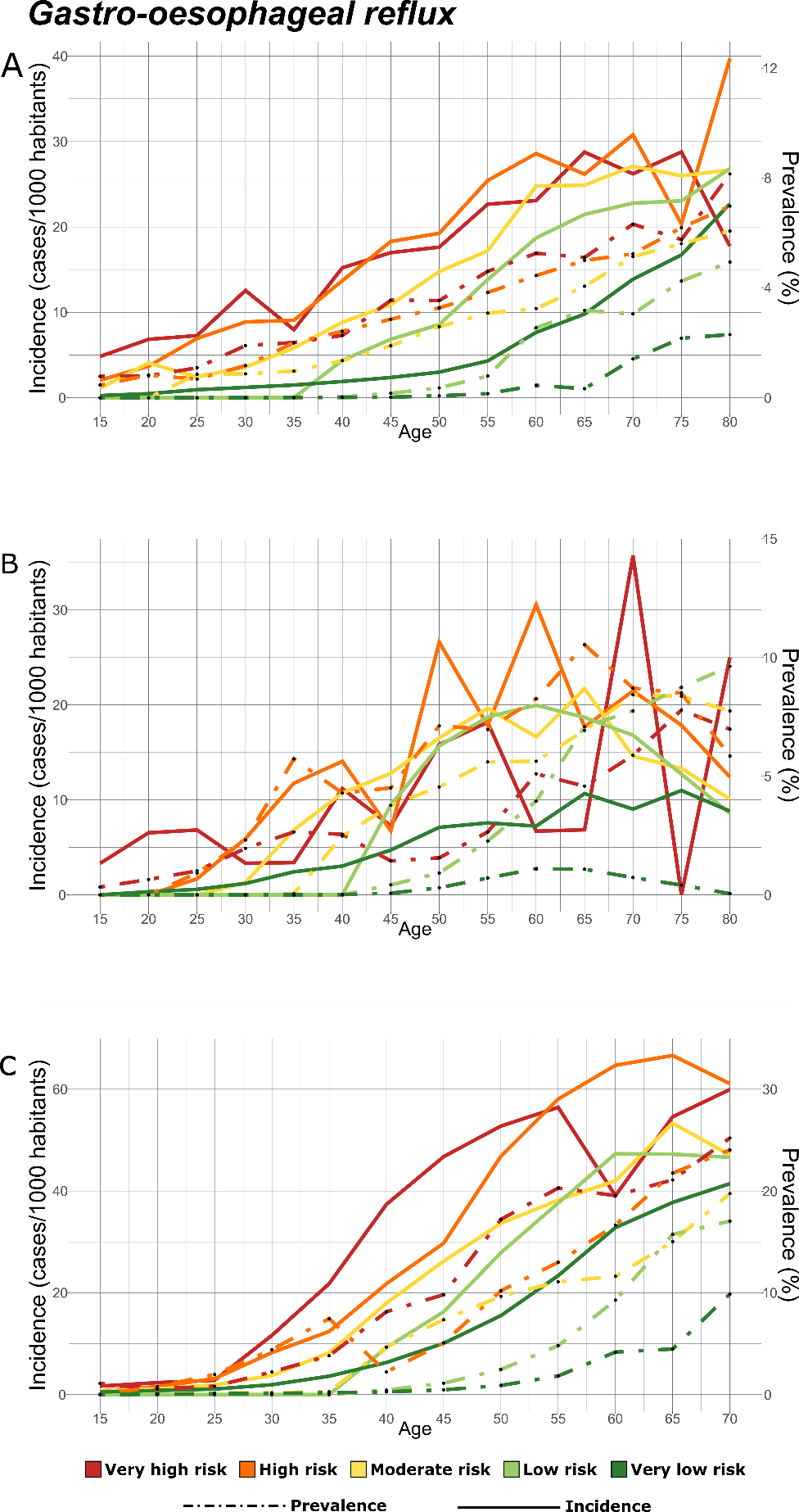

Figure S12 - Longitudinal analysis of disease prevalence and incidence of new onsets of gastro-oesophageal reflux (ICD-10-CM: K21). *Panel A) CHSS cohort; Panel B) THL cohort; Panel C)UKB cohort. Disease incidence is assessed in a 5-years interval, plotted in the left y-axis and represented with solid lines. Disease prevalence is plotted in the right y-axis and represented with dashed lines. The line colours correspond to the MADS risk pyramid tiers: red: very high-risk group; orange: high-risk group; yellow: medium-risk group; light green: low-risk group; green: very low-risk group.*

### Annex 3: Adjusted Disability Weights

| A/C | DW Acute | DW Chronic | ICD10 | Disease Name |
| --- | --- | --- | --- | --- |
| A | 0.0497 | - | D64 | Other anemias |
| C | - | 0.0190 | E03 | Other hypothyroidism |
| C | - | 0.0388 | E66 | Overweight and obesity |
| C | - | 0.0388 | E78 | Disorders of lipoprotein metabolism and other lipidemias |
| C | - | 0.1071 | F10 | Alcohol related disorders |
| C | - | 0.1414 | F17 | Nicotine dependence |
| A/C | 0.7780 | 0.5880 | F20 | Schizophrenia |
| A/C | 0.4920 | 0.0320 | F31 | Bipolar disorder |
| C | - | 0.2187 | F32 | Major depressive disorder, single episode |
| C | - | 0.2187 | F33 | Major depressive disorder, recurrent |
| C | - | 0.1036 | F40 | Phobic anxiety disorders |
| C | - | 0.1036 | F41 | Other anxiety disorders |
| C | - | 0.1036 | F43 | Reaction to severe stress, and adjustment disorders |
| C | - | 0.1036 | F45 | Somatoform disorders |
| C | - | 0.4942 | G40 | Epilepsy and recurrent seizures |
| C | - | 0.4410 | G43 | Migraine |
| A | 0.0100 | - | G47 | Sleep disorders |
| C | - | 0.0100 | G56 | Mononeuropathies of upper limb |
| A | 0.0702 | - | H10 | Conjunctivitis |
| A | 0.0702 | - | H53 | Visual disturbances |
| A/C | 0.0130 | 0.0032 | H65 | Nonsuppurative otitis media |
| A/C | 0.0130 | 0.0032 | H66 | Suppurative and unspecified otitis media |
| A | 0.1130 | - | H81 | Disorders of vestibular function |
| C | - | 0.1492 | H91 | Other and unspecified hearing loss |
| A | 0.1492 | - | H93 | Other disorders of ear, not elsewhere classified |
| C | - | 0.0871 | I10 | Essential (primary) hypertension |
| C | - | 0.0871 | I49 | Other cardiac arrhythmias |
| C | - | 0.4320 | I63 | Cerebral infarction |
| C | - | 0.4320 | I69 | Sequelae of cerebrovascular disease |
| A | 0.0871 | - | I83 | Varicose veins of lower extremities |
| A | 0.0871 | - | I95 | Hypotension |
| A | 0.0258 | - | J00 | Acute nasopharyngitis [common cold] |
| A | 0.0258 | - | J01 | Acute sinusitis |
| A | 0.0258 | - | J03 | Acute tonsillitis |
| A | 0.0258 | - | J06 | Acute upper respiratory infections of multiple and unspecified sites |
| A | 0.0633 | - | J18 | Pneumonia, unspecified organism |
| A | 0.0633 | - | J20 | Acute bronchitis |
| C | - | 0.0808 | J30 | Vasomotor and allergic rhinitis |
| C | - | 0.0808 | J31 | Chronic rhinitis, nasopharyngitis and pharyngitis |
| C | - | 0.2673 | J44 | Other chronic obstructive pulmonary disease |
| C | - | 0.0414 | J45 | Asthma |
| C | - | 0.0298 | K21 | Gastro-esophageal reflux disease |
| A | 0.3240 | - | K29 | Gastritis and duodenitis |
| A | 0.3240 | - | K35 | Acute appendicitis |
| A | 0.3240 | - | K43 | Ventral hernia |
| A | 0.3240 | - | K44 | Diaphragmatic hernia |
| A | 0.2310 | - | K52 | Other and unspecified noninfective gastroenteritis and colitis |
| A | 0.3240 | - | K56 | Paralytic ileus and intestinal obstruction without hernia |
| C | - | 0.0100 | K58 | Irritable bowel syndrome |
| A | 0.0100 | - | K59 | Other functional intestinal disorders |
| A | 0.5010 | - | K60 | Fissure and fistula of anal and rectal regions |
| A | 0.0100 | - | K62 | Other diseases of anus and rectum |
| C | - | 0.1780 | K76 | Other diseases of liver |
| A | 0.3240 | - | K80 | Cholelithiasis |
| C | - | 0.2637 | L20 | Atopic dermatitis |
| A | 0.0110 | - | L29 | Pruritus |
| A | 0.0110 | - | L30 | Other and unspecified dermatitis |
| C | - | 0.2637 | L40 | Psoriasis |
| A | 0.0914 | - | L50 | Urticaria |
| C | - | 0.0110 | L57 | Skin changes due to chronic exposure to nonionizing radiation |
| A | 0.0110 | - | L72 | Follicular cysts of skin and subcutaneous tissue |
| A | 0.0110 | - | L82 | Seborrheic keratosis |
| A | 0.0110 | - | L98 | Other disorders of skin and subcutaneous tissue, not elsewhere classified |
| C | - | 0.2472 | M06 | Other rheumatoid arthritis |
| C | - | 0.2950 | M10 | Gout |
| C | - | 0.0380 | M15 | Polyosteoarthritis |
| C | - | 0.0380 | M17 | Osteoarthritis of knee |
| C | - | 0.1700 | M20 | Acquired deformities of fingers and toes |
| C | - | 0.1700 | M24 | Other specific joint derangements |
| C | - | 0.1700 | M35 | Other systemic involvement of connective tissue |
| C | - | 0.1260 | M48 | Other spondylopathies |
| C | - | 0.1260 | M51 | Thoracic, thoracolumbar, and lumbosacral intervertebral disc disorders |
| A | 0.1064 | - | M54 | Dorsalgia |
| A | 0.1700 | - | M72 | Fibroblastic disorders |
| A | 0.1700 | - | M75 | Shoulder lesions |
| A | 0.1700 | - | M77 | Other enthesopathies |
| A | 0.1700 | - | M79 | Other and unspecified soft tissue disorders, not elsewhere classified |
| A | 0.0775 | - | N17 | Acute kidney failure |
| C | - | 0.3375 | N18 | Chronic kidney disease (CKD) |
| A | 0.0347 | - | N30 | Cystitis |
| C | - | 0.0670 | N40 | Benign prostatic hyperplasia |
| A | 0.0110 | - | N64 | Other disorders of breast |
| A | 0.0110 | - | N76 | Other inflammation of vagina and vulva |
| C | - | 0.0308 | N81 | Female genital prolapse |
| A | 0.0110 | - | N84 | Polyp of female genital tract |
| C | - | 0.0110 | N92 | Excessive, frequent and irregular menstruation |
| A | 0.0110 | - | N94 | Pain and other conditions associated with female genital organs and menstrual cycle |
| C | - | 0.0110 | N95 | Menopausal and other perimenopausal disorders |

### Annex 4: Probabilities of Relevance

| Disease | Interval | Onset Interval | PR |
| --- | --- | --- | --- |
| D64 | 1 | 1 | 0.207629 |
| E03 | 1 | 1 | 0.490952 |
| E66 | 1 | 1 | 0.514122 |
| E78 | 1 | 1 | 0.242673 |
| F10 | 1 | 1 | 0.537501 |
| F17 | 1 | 1 | 0.538888 |
| F41 | 1 | 1 | 1 |
| F43 | 1 | 1 | 0.996442 |
| G40 | 1 | 1 | 0.357502 |
| G43 | 1 | 1 | 0.587172 |
| G56 | 1 | 1 | 0.241626 |
| H10 | 1 | 1 | 0.281522 |
| H53 | 1 | 1 | 0.538474 |
| H65 | 1 | 1 | 0.537491 |
| H66 | 1 | 1 | 0.487149 |
| H81 | 1 | 1 | 0.217475 |
| H91 | 1 | 1 | 0.168141 |
| H93 | 1 | 1 | 0.158599 |
| I10 | 1 | 1 | 0.211192 |
| I49 | 1 | 1 | 0.222648 |
| I63 | 1 | 1 | 0.260996 |
| I83 | 1 | 1 | 0.223732 |
| I95 | 1 | 1 | 0.306704 |
| J00 | 1 | 1 | 0.257687 |
| J01 | 1 | 1 | 0.546747 |
| J03 | 1 | 1 | 0.72973 |
| J06 | 1 | 1 | 0.584992 |
| J18 | 1 | 1 | 0.482417 |
| J20 | 1 | 1 | 0.471248 |
| J30 | 1 | 1 | 0.703677 |
| J44 | 1 | 1 | 0.317264 |
| J45 | 1 | 1 | 0.750012 |
| K21 | 1 | 1 | 0.426771 |
| K29 | 1 | 1 | 0.21998 |
| K43 | 1 | 1 | 0.460557 |
| K44 | 1 | 1 | 0.244155 |
| K52 | 1 | 1 | 0.22367 |
| K56 | 1 | 1 | 0.179363 |
| K58 | 1 | 1 | 0.409958 |
| K59 | 1 | 1 | 0.23038 |
| K60 | 1 | 1 | 0.181343 |
| K62 | 1 | 1 | 0.232775 |
| K76 | 1 | 1 | 0.301806 |
| K80 | 1 | 1 | 0.236866 |
| L20 | 1 | 1 | 0.185839 |
| L29 | 1 | 1 | 0.278717 |
| L30 | 1 | 1 | 0.548071 |
| L40 | 1 | 1 | 0.096611 |
| L50 | 1 | 1 | 0.339331 |
| L57 | 1 | 1 | 0.275775 |
| L72 | 1 | 1 | 0.22363 |
| L82 | 1 | 1 | 0.289739 |
| L98 | 1 | 1 | 0.193764 |
| M10 | 1 | 1 | 0.533441 |
| M15 | 1 | 1 | 0.380628 |
| M17 | 1 | 1 | 0.223994 |
| M20 | 1 | 1 | 0.242078 |
| M48 | 1 | 1 | 0.185253 |
| M51 | 1 | 1 | 0.153944 |
| M54 | 1 | 1 | 0.358052 |
| M72 | 1 | 1 | 0.216581 |
| M75 | 1 | 1 | 0.382661 |
| M77 | 1 | 1 | 0.328523 |
| M79 | 1 | 1 | 0.676191 |
| N17 | 1 | 1 | 0.299834 |
| N18 | 1 | 1 | 0.250667 |
| N30 | 1 | 1 | 0.427956 |
| N40 | 1 | 1 | 0.27723 |
| N64 | 1 | 1 | 0.61271 |
| N76 | 1 | 1 | 0.233089 |
| N92 | 1 | 1 | 0.198923 |
| N94 | 1 | 1 | 0.504627 |
| N95 | 1 | 1 | 0.314981 |
| F20 | 1 | 1 | 1 |
| F31 | 1 | 1 | 0.617285 |
| F40 | 1 | 1 | 0.996332 |
| F45 | 1 | 1 | 0.191534 |
| G47 | 1 | 1 | 0.431321 |
| I69 | 1 | 1 | 0.105805 |
| J31 | 1 | 1 | 0.482109 |
| K35 | 1 | 1 | 0.128864 |
| M06 | 1 | 1 | 0.15675 |
| M24 | 1 | 1 | 0.268155 |
| M35 | 1 | 1 | 0.361547 |
| N81 | 1 | 1 | 0.33795 |
| N84 | 1 | 1 | 0.273644 |
| D64 | 2 | 2 | 0.145307 |
| E03 | 2 | 2 | 0.666467 |
| E66 | 2 | 2 | 0.90324 |
| E78 | 2 | 2 | 0.607976 |
| F10 | 2 | 2 | 0.932132 |
| F17 | 2 | 2 | 0.679072 |
| F41 | 2 | 2 | 0.666667 |
| F43 | 2 | 2 | 1 |
| G40 | 2 | 2 | 0.069491 |
| G43 | 2 | 2 | 0.522212 |
| G56 | 2 | 2 | 0.002524 |
| H10 | 2 | 2 | 0.177709 |
| H53 | 2 | 2 | 0.369519 |
| H65 | 2 | 2 | 0.088946 |
| H66 | 2 | 2 | 0.333856 |
| H81 | 2 | 2 | 0.001532 |
| H91 | 2 | 2 | 0.273224 |
| H93 | 2 | 2 | 0.08316 |
| I10 | 2 | 2 | 0.313948 |
| I49 | 2 | 2 | 0.011699 |
| I63 | 2 | 2 | 0.025087 |
| I83 | 2 | 2 | 0.004941 |
| I95 | 2 | 2 | 0.36688 |
| J00 | 2 | 2 | 0.255248 |
| J01 | 2 | 2 | 0.593139 |
| J03 | 2 | 2 | 0.30154 |
| J06 | 2 | 2 | 0.750867 |
| J18 | 2 | 2 | 0.359907 |
| J20 | 2 | 2 | 0.337761 |
| J30 | 2 | 2 | 0.664874 |
| J44 | 2 | 2 | 0.016355 |
| J45 | 2 | 2 | 0.58681 |
| K21 | 2 | 2 | 0.725603 |
| K29 | 2 | 2 | 0.216038 |
| K43 | 2 | 2 | 0.006751 |
| K44 | 2 | 2 | 0.007032 |
| K52 | 2 | 2 | 0.094769 |
| K56 | 2 | 2 | 0.037373 |
| K58 | 2 | 2 | 0.339967 |
| K59 | 2 | 2 | 0.019506 |
| K60 | 2 | 2 | 0.269061 |
| K62 | 2 | 2 | 0.118576 |
| K76 | 2 | 2 | 0.008777 |
| K80 | 2 | 2 | 0.00885 |
| L20 | 2 | 2 | 0.029375 |
| L29 | 2 | 2 | 0.077698 |
| L30 | 2 | 2 | 0.321978 |
| L40 | 2 | 2 | 0.36384 |
| L50 | 2 | 2 | 0.004706 |
| L57 | 2 | 2 | 0.014397 |
| L72 | 2 | 2 | 0.002464 |
| L82 | 2 | 2 | 0.025031 |
| L98 | 2 | 2 | 0.004672 |
| M10 | 2 | 2 | 0.229375 |
| M15 | 2 | 2 | 0.021844 |
| M17 | 2 | 2 | 0.008602 |
| M20 | 2 | 2 | 0.025575 |
| M48 | 2 | 2 | 0.543157 |
| M51 | 2 | 2 | 0.669599 |
| M54 | 2 | 2 | 1 |
| M72 | 2 | 2 | 0.005597 |
| M75 | 2 | 2 | 0.266992 |
| M77 | 2 | 2 | 0.268155 |
| M79 | 2 | 2 | 0.950369 |
| N17 | 2 | 2 | 0.126441 |
| N18 | 2 | 2 | 0.229945 |
| N30 | 2 | 2 | 0.13409 |
| N40 | 2 | 2 | 0.021651 |
| N64 | 2 | 2 | 0.036268 |
| N76 | 2 | 2 | 0.321532 |
| N92 | 2 | 2 | 0.611377 |
| N94 | 2 | 2 | 0.40996 |
| N95 | 2 | 2 | 0.020229 |
| F20 | 2 | 2 | 1 |
| F31 | 2 | 2 | 1 |
| F40 | 2 | 2 | 1 |
| F45 | 2 | 2 | 0.070387 |
| G47 | 2 | 2 | 0.664282 |
| I69 | 2 | 2 | 0.023645 |
| J31 | 2 | 2 | 0.027975 |
| K35 | 2 | 2 | 0.021178 |
| M06 | 2 | 2 | 0.182826 |
| M24 | 2 | 2 | 0.267584 |
| M35 | 2 | 2 | 0.26637 |
| N81 | 2 | 2 | 0.00845 |
| N84 | 2 | 2 | 0.026969 |
| D64 | 2 | 1 | 0.020895 |
| E03 | 2 | 1 | 0.639846 |
| E66 | 2 | 1 | 0.197175 |
| E78 | 2 | 1 | 0.5 |
| F10 | 2 | 1 | 0.50732 |
| F17 | 2 | 1 | 0.406307 |
| F41 | 2 | 1 | 0.699404 |
| F43 | 2 | 1 | 0.902009 |
| G40 | 2 | 1 | 0.071184 |
| G43 | 2 | 1 | 0.330547 |
| G56 | 2 | 1 | 0.059231 |
| H10 | 2 | 1 | 0.228742 |
| H53 | 2 | 1 | 0.113624 |
| H65 | 2 | 1 | 0.029636 |
| H66 | 2 | 1 | 0.003778 |
| H81 | 2 | 1 | 0.027335 |
| H91 | 2 | 1 | 0.018025 |
| H93 | 2 | 1 | 0.00194 |
| I10 | 2 | 1 | 0.012804 |
| I49 | 2 | 1 | 0.014845 |
| I63 | 2 | 1 | 0.054555 |
| I83 | 2 | 1 | 0.02105 |
| I95 | 2 | 1 | 0.12384 |
| J00 | 2 | 1 | 0.118451 |
| J01 | 2 | 1 | 0.009064 |
| J03 | 2 | 1 | 0.037438 |
| J06 | 2 | 1 | 0.196419 |
| J18 | 2 | 1 | 0.174266 |
| J20 | 2 | 1 | 0.081901 |
| J30 | 2 | 1 | 0.036414 |
| J44 | 2 | 1 | 0.017101 |
| J45 | 2 | 1 | 0.407349 |
| K21 | 2 | 1 | 0.400123 |
| K29 | 2 | 1 | 0.037 |
| K43 | 2 | 1 | 0.021352 |
| K44 | 2 | 1 | 0.002154 |
| K52 | 2 | 1 | 0.035781 |
| K56 | 2 | 1 | 0.058305 |
| K58 | 2 | 1 | 0.143014 |
| K59 | 2 | 1 | 0.025896 |
| K60 | 2 | 1 | 0.096265 |
| K62 | 2 | 1 | 0.166295 |
| K76 | 2 | 1 | 0.012471 |
| K80 | 2 | 1 | 0.02605 |
| L20 | 2 | 1 | 0.04015 |
| L29 | 2 | 1 | 0.029133 |
| L30 | 2 | 1 | 0.058341 |
| L40 | 2 | 1 | 0.099776 |
| L50 | 2 | 1 | 0.012935 |
| L57 | 2 | 1 | 0.010156 |
| L72 | 2 | 1 | 0.004805 |
| L82 | 2 | 1 | 0.010051 |
| L98 | 2 | 1 | 0.018125 |
| M10 | 2 | 1 | 0.039846 |
| M15 | 2 | 1 | 0.021502 |
| M17 | 2 | 1 | 0.05786 |
| M20 | 2 | 1 | 0.01368 |
| M48 | 2 | 1 | 0.557425 |
| M51 | 2 | 1 | 0.417676 |
| M54 | 2 | 1 | 0.370174 |
| M72 | 2 | 1 | 0.019751 |
| M75 | 2 | 1 | 0.006166 |
| M77 | 2 | 1 | 0.00928 |
| M79 | 2 | 1 | 0.210014 |
| N17 | 2 | 1 | 0.012911 |
| N18 | 2 | 1 | 0.327588 |
| N30 | 2 | 1 | 0.088923 |
| N40 | 2 | 1 | 0.238988 |
| N64 | 2 | 1 | 0.02793 |
| N76 | 2 | 1 | 0.310994 |
| N92 | 2 | 1 | 0.080761 |
| N94 | 2 | 1 | 0.181871 |
| N95 | 2 | 1 | 0.006917 |
| F20 | 2 | 1 | 0.928062 |
| F31 | 2 | 1 | 0.666667 |
| F40 | 2 | 1 | 0.666667 |
| F45 | 2 | 1 | 0.028975 |
| I69 | 2 | 1 | 8.48E-05 |
| J31 | 2 | 1 | 0.036557 |
| K35 | 2 | 1 | 0.019644 |
| M06 | 2 | 1 | 0.017095 |
| M24 | 2 | 1 | 0.010196 |
| M35 | 2 | 1 | 0.287653 |
| N81 | 2 | 1 | 0.012432 |
| N84 | 2 | 1 | 0.005068 |
| D64 | 3 | 3 | 0.000233 |
| E03 | 3 | 3 | 0.581504 |
| E66 | 3 | 3 | 0.215081 |
| E78 | 3 | 3 | 0.641215 |
| F10 | 3 | 3 | 0.655622 |
| F17 | 3 | 3 | 0.337533 |
| F41 | 3 | 3 | 1 |
| F43 | 3 | 3 | 1 |
| G40 | 3 | 3 | 7.64E-05 |
| G43 | 3 | 3 | 0.127064 |
| G56 | 3 | 3 | 0.02573 |
| H10 | 3 | 3 | 0.033825 |
| H53 | 3 | 3 | 0.003988 |
| H65 | 3 | 3 | 0.002786 |
| H66 | 3 | 3 | 0.336791 |
| H81 | 3 | 3 | 0.049376 |
| H91 | 3 | 3 | 0.006947 |
| H93 | 3 | 3 | 0.046048 |
| I10 | 3 | 3 | 0.666667 |
| I49 | 3 | 3 | 0.258034 |
| I63 | 3 | 3 | 0.000847 |
| I83 | 3 | 3 | 0.154023 |
| I95 | 3 | 3 | 0.006157 |
| J00 | 3 | 3 | 0.335306 |
| J01 | 3 | 3 | 0.153879 |
| J03 | 3 | 3 | 0.001042 |
| J06 | 3 | 3 | 0.333746 |
| J18 | 3 | 3 | 0.00034 |
| J20 | 3 | 3 | 0.048305 |
| J30 | 3 | 3 | 0.067269 |
| J44 | 3 | 3 | 0.200251 |
| J45 | 3 | 3 | 0.134008 |
| K21 | 3 | 3 | 0.804697 |
| K29 | 3 | 3 | 0.489027 |
| K43 | 3 | 3 | 0.000771 |
| K44 | 3 | 3 | 0.133885 |
| K52 | 3 | 3 | 0.437633 |
| K56 | 3 | 3 | 0.023764 |
| K58 | 3 | 3 | 0.838628 |
| K59 | 3 | 3 | 0.466593 |
| K60 | 3 | 3 | 0.00051 |
| K62 | 3 | 3 | 0.131354 |
| K76 | 3 | 3 | 0.003054 |
| K80 | 3 | 3 | 6.96E-05 |
| L20 | 3 | 3 | 0.014986 |
| L29 | 3 | 3 | 0.244383 |
| L30 | 3 | 3 | 0.079878 |
| L40 | 3 | 3 | 0.001048 |
| L50 | 3 | 3 | 0.000772 |
| L57 | 3 | 3 | 0.002291 |
| L72 | 3 | 3 | 0.002304 |
| L82 | 3 | 3 | 0.014331 |
| L98 | 3 | 3 | 0.043954 |
| M10 | 3 | 3 | 0.001691 |
| M15 | 3 | 3 | 0.01642 |
| M17 | 3 | 3 | 3.03E-05 |
| M20 | 3 | 3 | 0.001621 |
| M48 | 3 | 3 | 0.088179 |
| M51 | 3 | 3 | 0.139584 |
| M54 | 3 | 3 | 0.953653 |
| M72 | 3 | 3 | 0.002093 |
| M75 | 3 | 3 | 0.580773 |
| M77 | 3 | 3 | 0.100945 |
| M79 | 3 | 3 | 0.784598 |
| N17 | 3 | 3 | 0.027212 |
| N18 | 3 | 3 | 0.062883 |
| N30 | 3 | 3 | 0.327674 |
| N40 | 3 | 3 | 0.349648 |
| N64 | 3 | 3 | 0.0568 |
| N76 | 3 | 3 | 0.004811 |
| N92 | 3 | 3 | 0.03493 |
| N94 | 3 | 3 | 0.001314 |
| N95 | 3 | 3 | 0.279447 |
| F20 | 3 | 3 | 0.666667 |
| F31 | 3 | 3 | 1 |
| F40 | 3 | 3 | 0.336234 |
| F45 | 3 | 3 | 0.632903 |
| G47 | 3 | 3 | 0.666667 |
| I69 | 3 | 3 | 0.029474 |
| J31 | 3 | 3 | 0.001198 |
| K35 | 3 | 3 | 0.000174 |
| M06 | 3 | 3 | 0.002286 |
| M24 | 3 | 3 | 0.002296 |
| M35 | 3 | 3 | 0.334373 |
| N81 | 3 | 3 | 0.005425 |
| N84 | 3 | 3 | 0.001203 |
| D64 | 3 | 2 | 0.009784 |
| E03 | 3 | 2 | 0.581504 |
| E66 | 3 | 2 | 0.083618 |
| E78 | 3 | 2 | 0.210063 |
| F10 | 3 | 2 | 0.633333 |
| F17 | 3 | 2 | 0.099121 |
| F41 | 3 | 2 | 0.56674 |
| F43 | 3 | 2 | 0.600073 |
| G40 | 3 | 2 | 0.000487 |
| G43 | 3 | 2 | 0.133108 |
| G56 | 3 | 2 | 0.002473 |
| H10 | 3 | 2 | 0.011445 |
| H53 | 3 | 2 | 0.01428 |
| H65 | 3 | 2 | 0.016838 |
| H66 | 3 | 2 | 0.005995 |
| H81 | 3 | 2 | 0.010243 |
| H91 | 3 | 2 | 0.136588 |
| H93 | 3 | 2 | 4.27E-05 |
| I10 | 3 | 2 | 0.101343 |
| I49 | 3 | 2 | 0.011096 |
| I63 | 3 | 2 | 0.01957 |
| I83 | 3 | 2 | 0.005257 |
| I95 | 3 | 2 | 0.015927 |
| J00 | 3 | 2 | 0.002233 |
| J01 | 3 | 2 | 0.03338 |
| J03 | 3 | 2 | 0.004185 |
| J06 | 3 | 2 | 0.054189 |
| J18 | 3 | 2 | 0.004189 |
| J20 | 3 | 2 | 0.014529 |
| J30 | 3 | 2 | 0.002377 |
| J44 | 3 | 2 | 0.149343 |
| J45 | 3 | 2 | 0.234611 |
| K21 | 3 | 2 | 0.44033 |
| K29 | 3 | 2 | 0.070094 |
| K43 | 3 | 2 | 0.00893 |
| K44 | 3 | 2 | 0.114697 |
| K52 | 3 | 2 | 0.12342 |
| K56 | 3 | 2 | 0.006584 |
| K58 | 3 | 2 | 0.142484 |
| K59 | 3 | 2 | 0.466605 |
| K60 | 3 | 2 | 0.011556 |
| K62 | 3 | 2 | 0.041986 |
| K76 | 3 | 2 | 0.009094 |
| K80 | 3 | 2 | 0.000283 |
| L20 | 3 | 2 | 0.006752 |
| L29 | 3 | 2 | 0.024105 |
| L30 | 3 | 2 | 0.040511 |
| L40 | 3 | 2 | 0.013543 |
| L50 | 3 | 2 | 0.00626 |
| L57 | 3 | 2 | 0.000322 |
| L72 | 3 | 2 | 0.012774 |
| L82 | 3 | 2 | 0.000152 |
| L98 | 3 | 2 | 0.010782 |
| M10 | 3 | 2 | 0.025132 |
| M15 | 3 | 2 | 0.03191 |
| M17 | 3 | 2 | 0.010026 |
| M20 | 3 | 2 | 0.026688 |
| M48 | 3 | 2 | 0.125639 |
| M51 | 3 | 2 | 0.079216 |
| M54 | 3 | 2 | 0.066833 |
| M72 | 3 | 2 | 0.008739 |
| M75 | 3 | 2 | 0.02658 |
| M77 | 3 | 2 | 0.020682 |
| M79 | 3 | 2 | 0.185461 |
| N17 | 3 | 2 | 0.030636 |
| N18 | 3 | 2 | 0.032062 |
| N30 | 3 | 2 | 0.0101 |
| N40 | 3 | 2 | 0.336266 |
| N64 | 3 | 2 | 0.041827 |
| N76 | 3 | 2 | 0.006632 |
| N92 | 3 | 2 | 0.034444 |
| N94 | 3 | 2 | 0.035663 |
| N95 | 3 | 2 | 0.127145 |
| F20 | 3 | 2 | 0.666667 |
| F31 | 3 | 2 | 0.666667 |
| F40 | 3 | 2 | 0.335923 |
| F45 | 3 | 2 | 0.299765 |
| I69 | 3 | 2 | 0.020813 |
| J31 | 3 | 2 | 0.053806 |
| K35 | 3 | 2 | 0.00056 |
| M06 | 3 | 2 | 0.008837 |
| M24 | 3 | 2 | 0.005525 |
| M35 | 3 | 2 | 0.00814 |
| N81 | 3 | 2 | 0.005645 |
| N84 | 3 | 2 | 0.013678 |
| D64 | 4 | 4 | 0.001988 |
| E03 | 4 | 4 | 6.69E-05 |
| E66 | 4 | 4 | 0.60924 |
| E78 | 4 | 4 | 0.334674 |
| F10 | 4 | 4 | 0.732118 |
| F17 | 4 | 4 | 0.089398 |
| F41 | 4 | 4 | 0.672721 |
| F43 | 4 | 4 | 0.665326 |
| G40 | 4 | 4 | 0.002705 |
| G43 | 4 | 4 | 0.002275 |
| G56 | 4 | 4 | 0.009524 |
| H10 | 4 | 4 | 0.004784 |
| H53 | 4 | 4 | 0.002745 |
| H65 | 4 | 4 | 0.015268 |
| H66 | 4 | 4 | 0.009448 |
| H81 | 4 | 4 | 0.001592 |
| H91 | 4 | 4 | 0.004356 |
| H93 | 4 | 4 | 0.019601 |
| I10 | 4 | 4 | 0.6761 |
| I49 | 4 | 4 | 0.190506 |
| I63 | 4 | 4 | 0.334625 |
| I83 | 4 | 4 | 4.58E-05 |
| I95 | 4 | 4 | 0.003859 |
| J00 | 4 | 4 | 0.012023 |
| J01 | 4 | 4 | 0.001793 |
| J03 | 4 | 4 | 0.015985 |
| J06 | 4 | 4 | 0.482069 |
| J18 | 4 | 4 | 0.192174 |
| J20 | 4 | 4 | 0.002659 |
| J30 | 4 | 4 | 0.000795 |
| J44 | 4 | 4 | 0.074403 |
| J45 | 4 | 4 | 0.000279 |
| K21 | 4 | 4 | 0.100767 |
| K29 | 4 | 4 | 0.205315 |
| K43 | 4 | 4 | 0.004221 |
| K44 | 4 | 4 | 0.00207 |
| K52 | 4 | 4 | 0.001679 |
| K56 | 4 | 4 | 0.302046 |
| K58 | 4 | 4 | 0.108629 |
| K59 | 4 | 4 | 0.589787 |
| K60 | 4 | 4 | 0.00122 |
| K62 | 4 | 4 | 0.001992 |
| K76 | 4 | 4 | 0.292126 |
| K80 | 4 | 4 | 4.49E-06 |
| L20 | 4 | 4 | 0.000321 |
| L29 | 4 | 4 | 0.00341 |
| L30 | 4 | 4 | 0.002894 |
| L40 | 4 | 4 | 0.003798 |
| L50 | 4 | 4 | 0.001382 |
| L57 | 4 | 4 | 0.002819 |
| L72 | 4 | 4 | 0.005499 |
| L82 | 4 | 4 | 0.004651 |
| L98 | 4 | 4 | 0.001031 |
| M10 | 4 | 4 | 0.30812 |
| M15 | 4 | 4 | 0.357456 |
| M17 | 4 | 4 | 0.000121 |
| M20 | 4 | 4 | 0.001677 |
| M48 | 4 | 4 | 0.33424 |
| M51 | 4 | 4 | 0.024462 |
| M54 | 4 | 4 | 0.757775 |
| M72 | 4 | 4 | 0.003851 |
| M75 | 4 | 4 | 0.000228 |
| M77 | 4 | 4 | 0.001568 |
| M79 | 4 | 4 | 0.341395 |
| N17 | 4 | 4 | 0.074247 |
| N18 | 4 | 4 | 0.067345 |
| N30 | 4 | 4 | 0.334153 |
| N40 | 4 | 4 | 0.00023 |
| N64 | 4 | 4 | 0.011165 |
| N76 | 4 | 4 | 0.011783 |
| N92 | 4 | 4 | 0.006472 |
| N94 | 4 | 4 | 0.009572 |
| N95 | 4 | 4 | 0.100244 |
| F20 | 4 | 4 | 1 |
| F31 | 4 | 4 | 0.705556 |
| F40 | 4 | 4 | 0.018261 |
| F45 | 4 | 4 | 0.007279 |
| G47 | 4 | 4 | 0.334086 |
| I69 | 4 | 4 | 0.336072 |
| J31 | 4 | 4 | 0.007434 |
| K35 | 4 | 4 | 0.310043 |
| M06 | 4 | 4 | 0.001018 |
| M24 | 4 | 4 | 0.014846 |
| M35 | 4 | 4 | 0.008497 |
| N81 | 4 | 4 | 0.000114 |
| N84 | 4 | 4 | 0.001004 |
| D64 | 4 | 3 | 0.031453 |
| E03 | 4 | 3 | 1.28E-05 |
| E66 | 4 | 3 | 0.001795 |
| E78 | 4 | 3 | 0.002176 |
| F10 | 4 | 3 | 0.620467 |
| F17 | 4 | 3 | 0.066422 |
| F41 | 4 | 3 | 0.433664 |
| F43 | 4 | 3 | 0.843938 |
| G40 | 4 | 3 | 0.002973 |
| G43 | 4 | 3 | 0.007675 |
| G56 | 4 | 3 | 0.002237 |
| H10 | 4 | 3 | 0.030127 |
| H53 | 4 | 3 | 0.341754 |
| H65 | 4 | 3 | 0.023073 |
| H66 | 4 | 3 | 0.010569 |
| H81 | 4 | 3 | 0.00763 |
| H91 | 4 | 3 | 0.00385 |
| H93 | 4 | 3 | 0.0425 |
| I10 | 4 | 3 | 0.00061 |
| I49 | 4 | 3 | 0.006614 |
| I63 | 4 | 3 | 0.008991 |
| I83 | 4 | 3 | 0.000473 |
| I95 | 4 | 3 | 0.01024 |
| J00 | 4 | 3 | 0.018283 |
| J01 | 4 | 3 | 0.009354 |
| J03 | 4 | 3 | 0.037861 |
| J06 | 4 | 3 | 0.00273 |
| J18 | 4 | 3 | 0.008017 |
| J20 | 4 | 3 | 0.021369 |
| J30 | 4 | 3 | 0.003008 |
| J44 | 4 | 3 | 0.026563 |
| J45 | 4 | 3 | 0.001518 |
| K21 | 4 | 3 | 0.001271 |
| K29 | 4 | 3 | 0.002406 |
| K43 | 4 | 3 | 0.005458 |
| K44 | 4 | 3 | 0.029245 |
| K52 | 4 | 3 | 0.008246 |
| K56 | 4 | 3 | 0.01696 |
| K58 | 4 | 3 | 0.011087 |
| K59 | 4 | 3 | 0.075915 |
| K60 | 4 | 3 | 0.020382 |
| K62 | 4 | 3 | 0.002944 |
| K76 | 4 | 3 | 0.018396 |
| K80 | 4 | 3 | 0.00105 |
| L20 | 4 | 3 | 0.005096 |
| L29 | 4 | 3 | 0.007139 |
| L30 | 4 | 3 | 0.005837 |
| L40 | 4 | 3 | 0.005635 |
| L50 | 4 | 3 | 0.019405 |
| L57 | 4 | 3 | 0.015421 |
| L72 | 4 | 3 | 0.014275 |
| L82 | 4 | 3 | 0.011029 |
| L98 | 4 | 3 | 0.030898 |
| M10 | 4 | 3 | 0.010196 |
| M15 | 4 | 3 | 0.012109 |
| M17 | 4 | 3 | 0.006472 |
| M20 | 4 | 3 | 0.002886 |
| M48 | 4 | 3 | 0.336163 |
| M51 | 4 | 3 | 0.012603 |
| M54 | 4 | 3 | 0.231507 |
| M72 | 4 | 3 | 0.011313 |
| M75 | 4 | 3 | 0.204019 |
| M77 | 4 | 3 | 0.003613 |
| M79 | 4 | 3 | 0.026842 |
| N17 | 4 | 3 | 0.086403 |
| N18 | 4 | 3 | 0.031897 |
| N30 | 4 | 3 | 0.005342 |
| N40 | 4 | 3 | 0.001307 |
| N64 | 4 | 3 | 0.012489 |
| N76 | 4 | 3 | 0.025574 |
| N92 | 4 | 3 | 0.008984 |
| N94 | 4 | 3 | 0.009884 |
| N95 | 4 | 3 | 0.0008 |
| F20 | 4 | 3 | 0.770102 |
| F31 | 4 | 3 | 0.706754 |
| F40 | 4 | 3 | 0.036717 |
| F45 | 4 | 3 | 0.00867 |
| G47 | 4 | 3 | 0.284504 |
| I69 | 4 | 3 | 0.35072 |
| J31 | 4 | 3 | 0.011781 |
| K35 | 4 | 3 | 0.31619 |
| M06 | 4 | 3 | 0.007894 |
| M24 | 4 | 3 | 0.115725 |
| M35 | 4 | 3 | 0.018638 |
| N81 | 4 | 3 | 0.005471 |
| N84 | 4 | 3 | 0.002112 |
